# MedGenesis: Toward a World Model for Autonomous Clinical and Translational Research

**DOI:** 10.64898/2026.06.14.26355612

**Authors:** Hao Xiao, Nan Jiang, Tiancheng Zhang, Zhenfei Yin, Tao Gui, Zhongyue Zhang, Ke Shao, Jiacheng Ge, Rongyuan Wei, Jiaomeng Pan, Jiaqiang Ma, Ling Yang, Zhe Zhao, Jian Zhou, Jia Fan, Yugang Jiang, Philip Torr, Shuangjia Zheng, Yingcheng Wu, Qiang Gao

**Affiliations:** Department of Hepatobiliary Surgery & Transplantation, Liver Cancer Institute and State Key Laboratory of Genetics and Development of Complex Phenotypes, Zhongshan Hospital, Fudan University; Key Laboratory of Carcinogenesis & Cancer Invasion, Ministry of Education; Institutes of Biomedical Sciences; Human Phenome Institute, Fudan University; Shanghai Academy of Natural Sciences, Shanghai, China; Department of Engineering Science, University of Oxford, UK; College of Computer Science and Artificial Intelligence, Fudan University, Shanghai, China; School of Artificial Intelligence; Global Institute of Future Technology, Shanghai Jiao Tong University; Lingang Laboratory, Shanghai, China; NYU Grossman School of Medicine, New York University, New York, USA; Department of Electrical and Computer Engineering, Princeton AI Lab, Princeton University, New Jersey, USA; Department of Pathology, Department of Genetics, Stanford University School of Medicine, Stanford, CA, USA

**Keywords:** MedGenesis, clinical AI scientist, clinical world model, closed-loop clinical research, hypothesis-to-action reasoning, electronic health records, evidence synthesis, mechanistic validation

## Abstract

Clinical research advances slowly because its core tasks, from evidence synthesis to mechanistic validation, remain fragmented. We present MedGenesis, a clinical artificial intelligence (AI) scientist built on a world-model reasoning loop that jointly updates a Latent Hypothesis Space and a Latent Action Space under expected information gain (EIG), uncertainty reduction (UR), and a safety prior P(safe), and integrates longitudinal electronic health records (EHRs) via the Virtual Clinical Trajectory and Observation Representation (ViCTOR) for cohort retrieval, trajectory stratification, and time-to-event analysis. On two benchmarks—ClinicalResBench (1,697 expert-curated questions) and ClinicalRepBench (40 paper-reproduction tasks)—MedGenesis outperformed frontier language models and biomedical AI systems while reducing hallucination. Across 1 million patient observations spanning five clinical evidence formats, it generated traceable outputs across meta-analysis, randomized controlled trials, real-world trajectories, case-control studies, and case reports, with one wet-lab-coupled run nominating a 3-hydroxybutyrate– neutrophil axis modulating antitumor immunity. These results compress hypothesis-to-evidence cycles from years to hours, creating a continuous clinical discovery process.

## INTRODUCTION

Clinical research has entered a data-rich but progress-constrained era. Modern evidence-based medicine increasingly depends on real-world evidence, longitudinal patient trajectories, molecular profiling, and population-scale biomedical resources, rather than randomized trials alone^1^. Hospitals continuously generate EHRs, laboratory results, medication exposures, imaging and pathology reports, adverse-event records, and follow-up outcomes, while public resources provide clinical trials, population cohorts, multi-omics atlases, epidemiologic surveys, and biomedical literature^2,3^. In principle, these data should accelerate evidence synthesis, treatment-sub-group discovery, trial enrichment, pharmacovigilance, rare-case retrieval, and biomarker generation. In practice, however, data-to-progress conversion remains inefficient. Clinical data are irregular, incomplete, multimodal, and institution-specific, and molecular or population datasets are often disconnected from patient trajectories and clinical questions.

Consequently, clinically meaningful observations still require manual chart review, cohort definition, endpoint curation, statistical redesign, and expert interpretation before they become publishable evidence or follow-up experiments. The central bottleneck is therefore the lack of a reliable system that converts fragmented clinical and biomedical data into auditable hypotheses, executable analyses, and clinically meaningful next actions.

Recent agentic large language model (LLM) systems have begun to address fragments of this lifecycle. DeepRare and MAI-DxO match expert physicians on differential diagnosis^4,5^. OriGene nominates druggable targets validated in patient-derived organoids^6^. Biomni unifies hundreds of bioinformatics skills for end-to-end biological analysis^7^, and Google’s AI Co-Scientist evolves hypotheses through tournament-style multi-agent debate^8^. In parallel, peer-reviewed medical AI studies have shown rapid progress in expert-level medical question answering, simulated diagnostic dialogue, differential-diagnosis support, and multimodal biomedical foundation models^9-13^. These systems are important advances, but each remains anchored to a single research stage, such as diagnosis, target nomination, hypothesis brainstorming, or analysis automation. None has yet produced candidate outputs across the heterogeneous formats demanded by the clinical-research lifecycle, including Preferred Reporting Items for Systematic Reviews and Meta-Analyses (PRISMA)-compliant pooled estimates, trial-enrichment hypotheses, pharmacovigilance panels, risk-stratification scores, and mechanistic hypotheses supported by primary wet-lab data. Four methodological gaps remain. First, hypothesis generation is often decoupled from action selection. Second, outputs rarely undergo adversarial scrutiny for confounding, selection bias, leakage, and missing data, despite reported hallucination rates of 15–30% on clinical tasks^14,15^. Third, heterogeneous evidence formats cannot be collapsed into a single output template. Fourth, claims are not reliably traceable, so expert feedback cannot refine the system without retraining.

Addressing these gaps requires moving beyond stage-specific agents to a system that preserves continuity between a candidate claim, the evidence needed to test it, the action selected to obtain that evidence, and the boundary under which the claim can be reported. In clinical research, the useful output is therefore not a standalone answer, but a traceable evidence state that can be reviewed, revised, and advanced to the next study step.

Here, we present MedGenesis, a clinical AI scientist built on a Clinical World Model for closed-loop clinical research. MedGenesis links a Latent Hypothesis Space with a Latent Action Space governed by expected information gain (EIG), uncertainty reduction (UR), and a safety prior (P(safe)), pairing each candidate claim with a safe and informative next action rather than reporting isolated outputs. For longitudinal patient-data questions, MedGenesis incorporates the Virtual Clinical Trajectory and Observation Representation (ViCTOR) patient-state layer, allowing cohort retrieval, trajectory stratification, and time-to-event analysis to enter the same research loop. We evaluate MedGenesis across clinical-research benchmarks and representative tasks from evidence synthesis and trial enrichment to real-world analysis and wet-lab-supported mechanism discovery.

## RESULTS

### MedGenesis: A world model toward end-to-end clinical study

MedGenesis was designed to make each clinical discovery step conditional on the result of the previous one, rather than executing retrieval or predefined analyses in a single pass. To support this closed-loop process at population scale, MedGenesis accesses public biomedical resources, including The Cancer Genome Atlas^16^ (TCGA), Gene Expression Omnibus (GEO)^17^, Genotype-Tissue Expression (GTEx)^18^, Clinical Proteomic Tumor Analysis Consortium (CPTAC)^19^, PubMed^20^, Surveillance, Epidemiology, and End Results (SEER)^21^, and the National Health and Nutrition Examination Survey (NHANES)^22^. It also accesses controlled resources, including UK Biobank^23^ and Medical Information Mart for Intensive Care IV (MIMIC-IV)^24^, together with institutional clinical records from Zhongshan Hospital, Fudan University. The institutional resources include longitudinal EHRs, pathology and imaging archives, outcome and follow-up registries, and ViCTOR patient representations (Figure 1A). Together, these resources expose at least 13 biomedical and clinical resources, more than 1 million clinical observations, 103,812 ViCTOR patient embeddings, at least 8 data modalities, and over 20,000 dynamic derived features. Each resource is wrapped as a callable skill within the Skill Ocean (Figure S1A), allowing MedGenesis to assemble the relevant world state on demand for each hypothesis.

**Figure 1.**
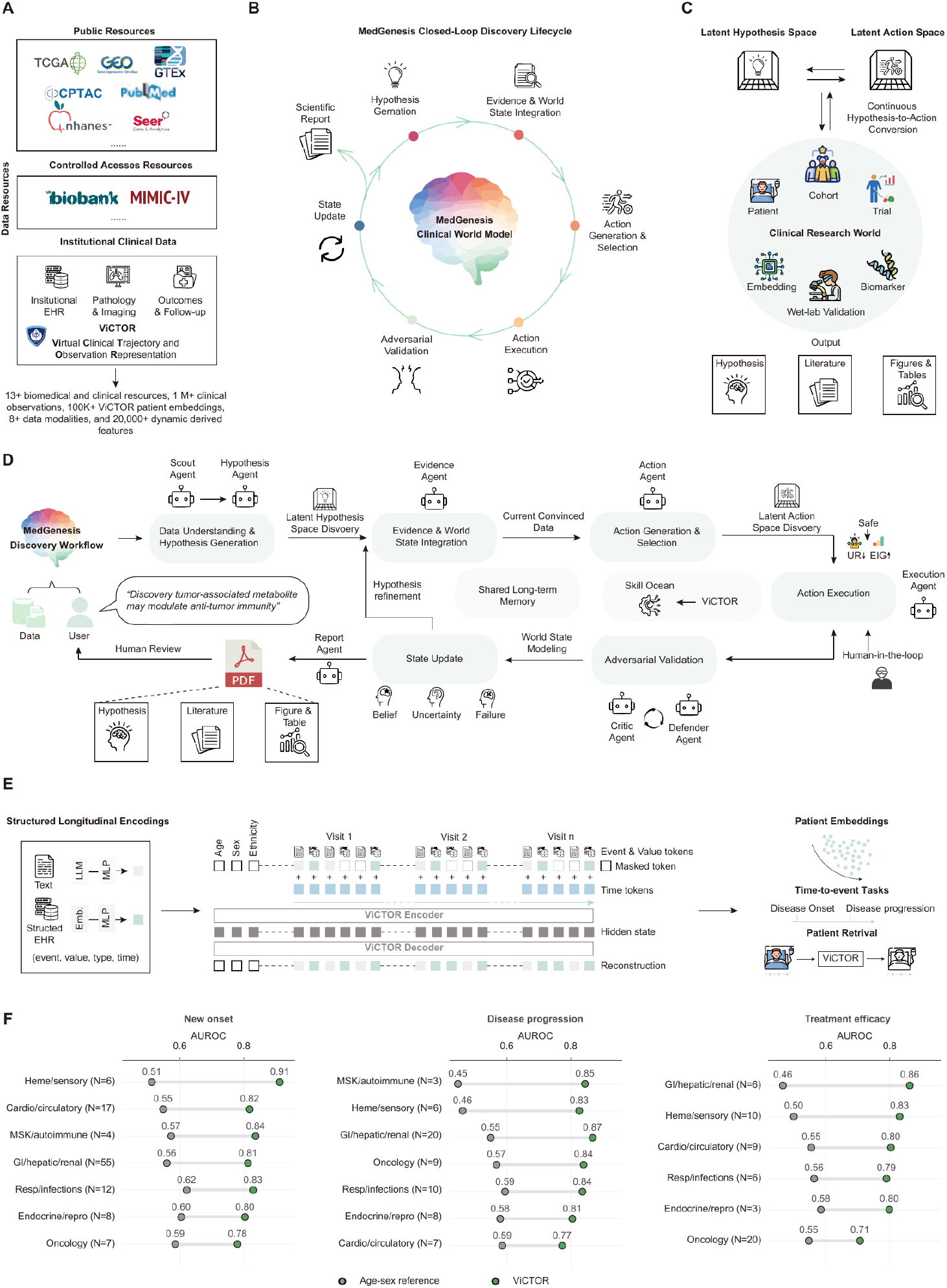
Architecture, data foundation, and patient-state layer of MedGenesis. (A) Data foundation underlying MedGenesis. Public resources include TCGA, GEO, GTEx, CPTAC, PubMed, NHANES, and SEER; controlled-access resources include UK Biobank and MIMIC-IV; and institutional resources from Zhongshan Hospital include longitudinal electronic health records (EHR), pathology and imaging archives, outcomes, follow-up registries, and ViCTOR-Zhongshan patient representations. The integrated foundation contains at least 13 biomedical and clinical resources, more than 1 million clinical observations, 103,812 ViCTOR patient embeddings, at least 8 data modalities, and more than 20,000 dynamic derived features. (B) Closed-loop discovery lifecycle of the MedGenesis Clinical World Model. Each cycle includes hypothesis generation, evidence and world-state integration, action generation and selection, action execution, adversarial validation, state update, and scientific report generation. (C) Continuous hypothesis-to-action conversion. The Latent Hypothesis Space and Latent Action Space are linked to the Clinical Research World, which includes patients, cohorts, trials, embeddings, biomarkers, and wet-lab validation. Outputs are rendered as hypotheses, literature summaries, and figures or tables. (D) End-to-end MedGenesis discovery workflow. From a user-supplied question and data inputs, Scout and Hypothesis Agents define candidate hypotheses, Evidence and Action Agents assemble the world state and rank actions, the Execution Agent calls Skill Ocean skills or human-in-the-loop wet-lab handoff, and Critic and Defender Agents perform adversarial validation before the Report Agent writes belief, uncertainty, and failure modes back into shared long-term memory. (E) ViCTOR-Zhongshan patient-state model. Structured EHR events and text-derived observations are encoded as event-value-time tokens across visits. The ViCTOR encoder-decoder reconstructs masked and temporal clinical states and generates patient embeddings for time-to-event prediction and patient retrieval. (F) ViCTOR time-to-event prediction benchmark. AUC distributions compare ViCTOR with an age-sex reference model across new-onset disease, disease-progression, and treatment-efficacy tasks grouped by clinical disease families. See also Figure S1. Performance metrics are presented as AUC distributions where shown.

The system operates through a six-stage lifecycle: Hypothesis Generation, Evidence and World-State Integration, Action Generation and Selection, Action Execution, Adversarial Validation, and State Update (Figure 1B, STAR Methods). At its core are two coupled spaces: a Latent Hypothesis Space indexed by plausibility and novelty, and a Latent Action Space indexed by EIG, UR, and P(safe) (Figure 1C). At each round, MedGenesis selects the action with the highest composite priority *α = EIG* · *UR* · *P(safe)*. It executes that action against the current Clinical Research World and uses the returned evidence to update both spaces. The next action is therefore chosen to reduce uncertainty among competing hypotheses, not simply to confirm the current leading explanation. This lifecycle is implemented by eight specialized agents operating over shared long-term memory and a dynamically extensible Skill Ocean (Figure 1D). The Scout and Hypothesis Agents define falsifiable claims. The Evidence and Action Agents assemble the relevant world state and select the next action. The Execution Agent runs the selected skill-mediated analysis or human-in-the-loop wet-lab handoff. The Critic, Defender, and Report Agents review the emerging claim before memory write-back. This governance layer keeps confounding, selection bias, leakage, unsupported inference, and residual uncertainty inside the discovery loop rather than relegating them to post hoc limitations.

To support hypothesis generation and action selection over real longitudinal patient data, MedGenesis incorporates ViCTOR as its patient-state modeling layer (Figure 1E, STAR Methods). ViCTOR converts structured clinical events and text-derived observations into longitudinal patient-state embeddings and is called by MedGenesis through the Skill Ocean for cohort retrieval, patient-state stratification, disease-trajectory analysis, virtual recruitment, and time-to-event modeling. Across 226 clinical prediction tasks, ViCTOR outperformed an age-sex reference model across disease families, including 109 new-onset disease tasks, 63 disease-progression tasks, and 54 treatment-efficacy tasks (Figure 1F). ViCTOR also improved disease-trajectory retrieval across 45 retrieval tasks spanning six disease families, where patient-state retrieval achieved higher top-five retrieval accuracy than retrieval based on the last progress note alone (Figure S1B). Together, these results position ViCTOR as the patient-state modeling layer of MedGenesis, linking closed-loop hypothesis-action reasoning to real longitudinal patient trajectories.

### MedGenesis outperforms frontier agents across clinical domains and study designs

A central difficulty in evaluating AI systems for clinical discovery is that most biomedical benchmarks test retrieval or single-turn question answering, whereas clinical research requires sustained reasoning across study designs, domain knowledge, and end-to-end reproduction of published findings. Recent systematic evaluation work has similarly emphasized that many healthcare large language model evaluations still rely on examination-style question answering rather than real patient-care data^25^. To probe MedGenesis along both axes, we constructed two complementary benchmarks for this study (Figure 2A). ClinicalResBench is a dual-track question-answering benchmark containing 1,697 question-answer pairs curated from clinical literature, guidelines, translational-medicine sources, and real-world evidence by a panel of domain experts with LLM-assisted drafting, quality control, ambiguity filtering, and final adjudication. It is split into ClinicalRes-Knowledge (1,256 questions over 10 clinical domains) and ClinicalRes-Methodology (441 questions over 5 study-design categories; Figure S2A). ClinicalRepBench takes the evaluation one step further: for each of 10 clinical domains, we selected 4 representative research papers and reconstructed them as 40 end-to-end reproduction tasks. Tasks were scored on a 0–100 scale, with 50 corresponding to human-expert level. The Re-Discovery / New-Discovery distinction separates reproducing a paper’s reported finding from identifying a previously unreported finding in the same data. We benchmarked MedGenesis against three classes of baselines: frontier LLMs (Gemini 3 Pro^26^, GPT-5.2^27^, Claude 4.5 Opus^28^, Grok 4.1^29^, DeepSeek-V3.2-Thinking^30^, Llama 4^31^), a domain-specific multi-agent system (Biomni), and two code-executing agents evaluated on ClinicalRepBench (Codex command-line interface (Codex CLI)^32^ and Claude Code^33^).

**Figure 2.**
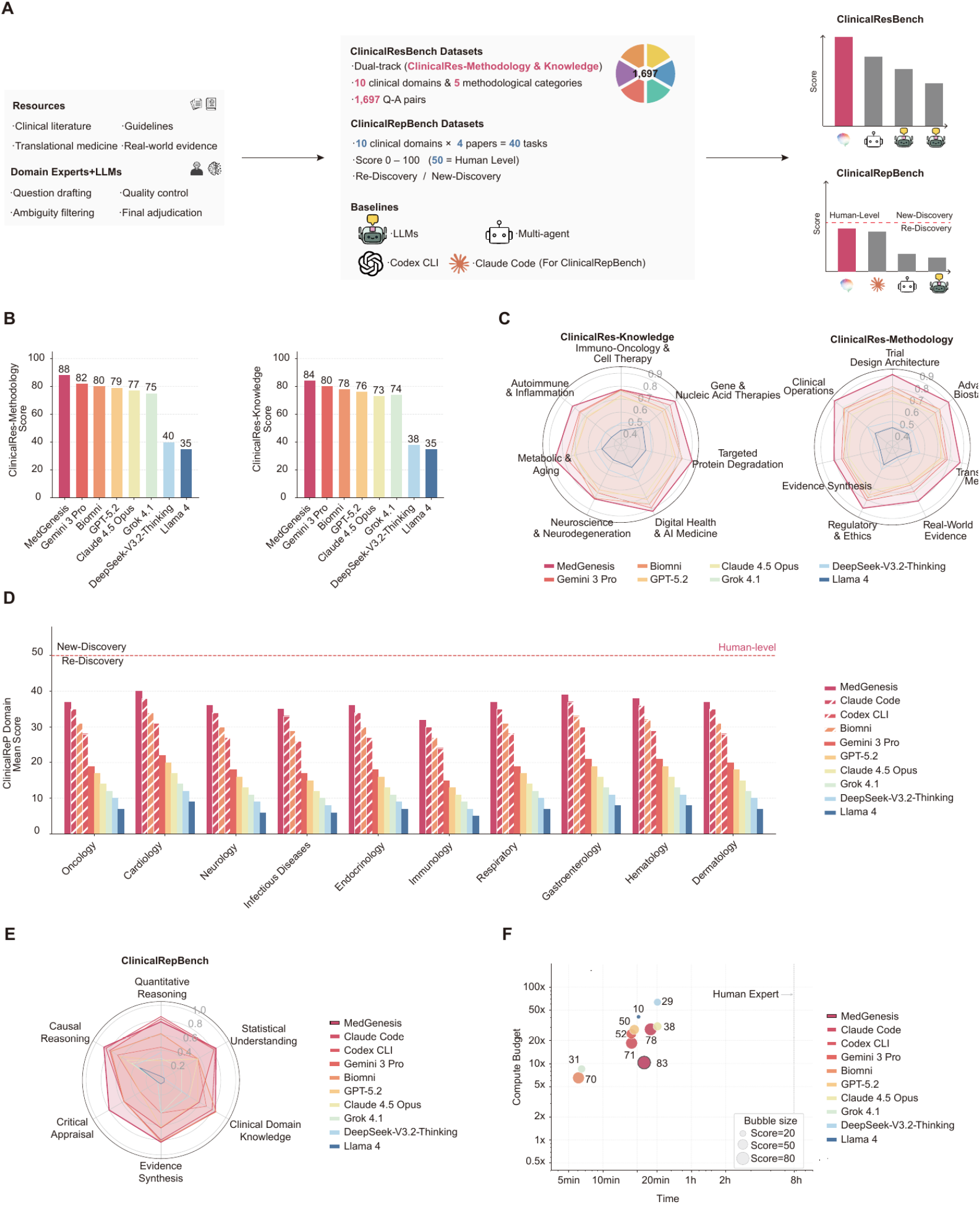
Benchmark construction and performance of MedGenesis across clinical domains and study designs. (A) Construction of ClinicalResBench and ClinicalRepBench. ClinicalResBench was built from clinical literature, guidelines, translational medicine, and real-world evidence with domain-expert and LLM-assisted question drafting, ambiguity filtering, quality control, and final adjudication. It contains 1,697 question-answer pairs across ClinicalRes-Knowledge and ClinicalRes-Methodology. ClinicalRepBench contains 40 paper-reproduction tasks across 10 clinical domains, scored from 0 to 100, with 50 representing the human-level reference threshold. (B) Overall ClinicalResBench scores for the methodology track (left) and knowledge track (right). MedGenesis achieved the highest scores on both tracks, reaching 88 on ClinicalRes-Methodology and 84 on ClinicalRes-Knowledge. (C) Domain- and methodology-level radar plots for ClinicalResBench. ClinicalRes-Knowledge covers immuno-oncology and cell therapy, gene and nucleic-acid therapies, targeted protein degradation, digital health and AI medicine, neuroscience and neurodegeneration, metabolic and aging, and autoimmune and inflammation. ClinicalRes-Methodology covers trial design architecture, advanced biostatistics, translational methods, real-world evidence, regulatory and ethics, evidence synthesis, and clinical operations. (D) ClinicalRepBench per-domain scores across oncology, cardiology, neurology, infectious diseases, endocrinology, immunology, respiratory medicine, gastroenterology, hematology, and dermatology. Dashed lines denote the Re-Discovery/New-Discovery boundary and the human-level reference threshold. (E) ClinicalRepBench capability radar. Six dimensions were evaluated: quantitative reasoning, statistical understanding, clinical-domain knowledge, evidence synthesis, critical appraisal, and causal reasoning. (F) Compute-budget and wall-clock-time efficiency frontier on ClinicalRepBench. Bubble size represents the composite score. MedGenesis reached the highest plotted score at moderate compute and time, approaching the human-expert reference while outperforming baseline systems under comparable or greater compute budgets. See also Figure S2.

On ClinicalResBench, MedGenesis achieved the top score on both tracks, reaching 88 on ClinicalRes-Methodology and 84 on ClinicalRes-Knowledge. It outperformed Gemini 3 Pro (82 and 80) and showed larger margins over Biomni, GPT-5.2, Claude 4.5 Opus, Grok 4.1, DeepSeek-V3.2-Thinking, and Llama 4 (Figure 2B). The performance margin was broad rather than limited to one question family. Across the domain and methodology axes shown in Figure 2C, and when recall was stratified by clinical domain and study-design category (Figure S2B), MedGenesis remained at the outer envelope of the comparison. The benchmark also covered a broad clinical-to-basic task space, spanning patient phenotyping, clinical narratives, structured EHRs, medical imaging, multi-omics data, biomedical knowledge, and molecular structures (Figure S2C). MedGenesis achieved higher recall across these categories, rather than performing well only on text-based clinical questions. It also showed clear test-time scaling. When the inference budget increased from 1× to 9×, ClinicalRes-Methodology recall rose from approximately 0.85 to 0.95, and ClinicalRes-Knowledge recall rose from approximately 0.82 to 0.92. By contrast, static LLM baselines changed little across the same compute range (Figure S2D). This scaling pattern indicates that MedGenesis uses additional compute for deeper hypothesis-action refinement, not merely for repeated sampling.

ClinicalRepBench was more demanding because it required end-to-end reproduction of published clinical studies. Across all 10 clinical domains, MedGenesis achieved a mean score of 32.40, outperforming the code-executing Claude Code and Codex CLI agents, the biomedical agent Biomni, and all frontier LLMs while remaining below the human-expert threshold of 50 (Figure 2D). This gap identifies the distance still to be closed. The radar view of six evaluation dimensions, including Quantitative Reasoning, Statistical Understanding, Clinical Domain Knowledge, Evidence Synthesis, Critical Appraisal, and Causal Reasoning, showed broad gains across capabilities, with particularly large margins on Causal Reasoning and Critical Appraisal (Figure 2E). To compare systems on accuracy, time, and compute jointly, we summarized each system’s ClinicalRepBench performance as the average of the six dimensions in Figure 2E multiplied by 10, yielding a single 0– 100 score. Under this summary, MedGenesis reached a score of 83 in approximately 20 minutes at a compute budget of about 10×. DeepSeek-V3.2-Thinking, Claude 4.5 Opus, and Codex CLI required 25–50× compute without matching this accuracy, and the human-expert reference point lay further to the right at approximately 8 hours (Figure 2F). Together, these results show that the closed-loop hypothesis-action architecture of MedGenesis improves both accuracy and the efficiency of clinical-discovery work at scale.

### MedGenesis autonomously generates hypotheses and actions throughout the discovery cycle

Benchmark scores indicate whether a system answers correctly, but not whether it behaves like a clinical scientist. We therefore evaluated the hypothesis-to-action behavior of MedGenesis using an 18-task clinical hypothesis-generation benchmark and an action-selection benchmark (STAR Methods). In the Latent Hypothesis Space, candidate claims were mapped by novelty and plausibility. Across five refinement rounds, MedGenesis moved from conservative, low-novelty starting claims toward the high-novelty, high-plausibility Ideal region (Figure 3A). The quantitative comparison confirmed this trajectory: MedGenesis was the only evaluated system with both novelty and plausibility concentrated in the predefined Ideal zone (≥0.75; Figure 3B). Claude Code and Codex CLI retained relatively high plausibility but generated less novel claims, whereas Biomni, Gemini 3 Pro, GPT-5.2, Claude 4.5 Opus, Grok 4.1, DeepSeek-V3.2-Thinking, and Llama 4 fell below the Ideal threshold on at least one axis. Representative matched tasks in metabolic medicine, oncology, and cardiology showed the same pattern (Figure S3).

**Figure 3.**
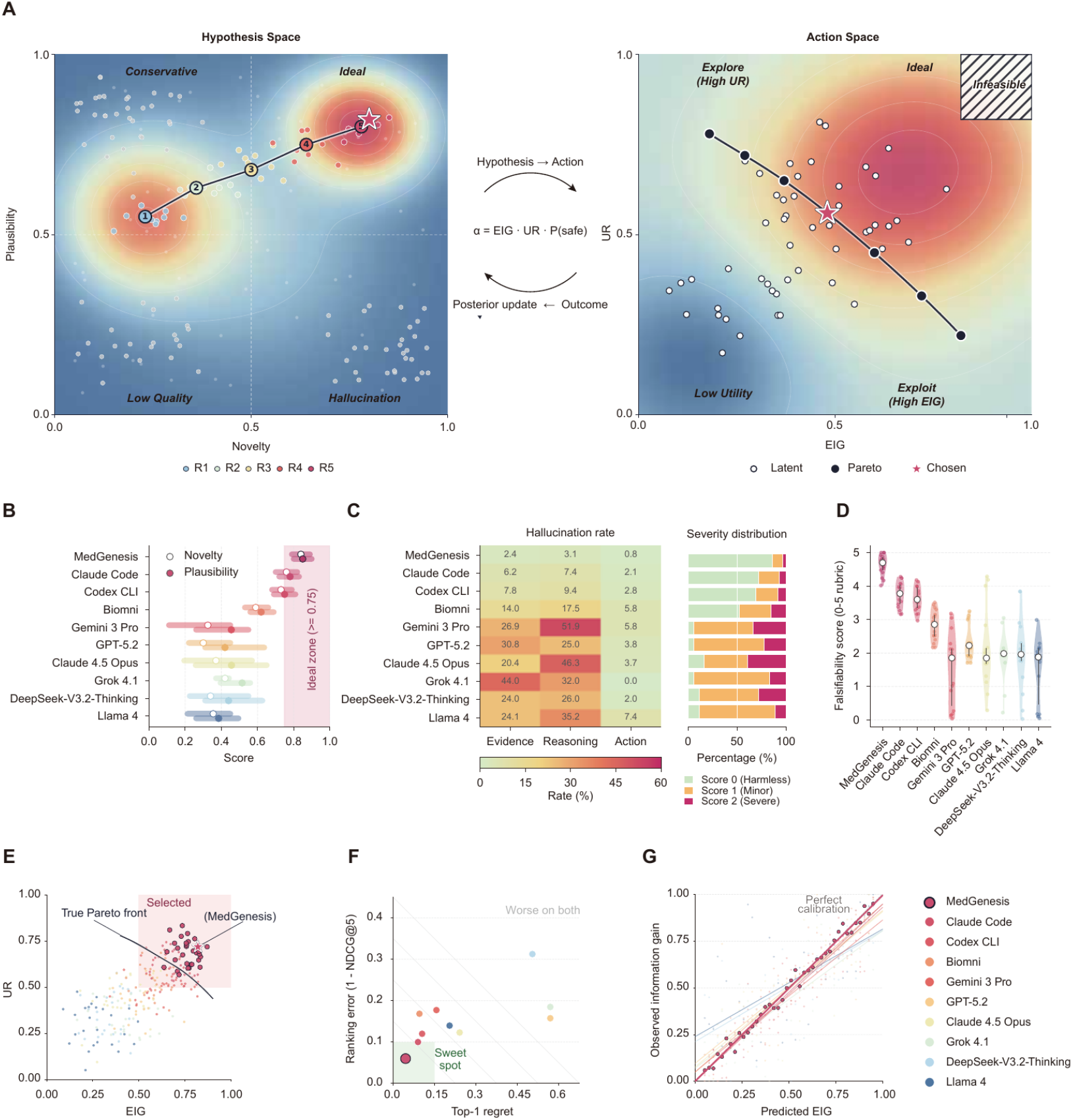
MedGenesis generates high-novelty, falsifiable, and calibrated hypotheses with low hallucination. (A) Joint hypothesis-action trajectory. Left: MedGenesis moves through the Latent Hypothesis Space, defined by novelty and plausibility, from conservative initial hypotheses to the high-novelty and high-plausibility Ideal region over five refinement rounds (R1-R5). Right: candidate actions are positioned in the Latent Action Space by expected information gain (EIG) and uncertainty reduction (UR). The selected MedGenesis action lies near the feasible Pareto front under the composite priority α = EIG × UR × P(safe). (B) Novelty and plausibility distributions across evaluated systems. Open circles denote novelty and filled circles denote plausibility; values are shown as median with interquartile range. The shaded Ideal zone indicates scores ≥0.75. (C) Hallucination rate and severity distribution by reasoning stage. Claim-level hallucinations were grouped as evidence, reasoning, or action errors. Severity was graded as Score 0 (harmless), Score 1 (minor), or Score 2 (severe). (D) Falsifiability score for generated hypotheses. Scores were assigned on a 0–5 rubric based on explicit mechanism, direction of effect, measurable decision boundary, testable cohort or endpoint, and refutation condition. (E) Action-selection quality in the EIG-UR space. Candidate actions are shown with the true Pareto front and the MedGenesis-selected region. MedGenesis-selected actions are enriched in the high-EIG and high-UR region near the Pareto front. (F) Portfolio action-ranking benchmark. Systems were compared by ranking error, defined as 1 − NDCG@5, and top-1 regret across pooled candidate action portfolios. Lower values indicate better prioritization of high-utility, executable, and safe actions. (G) Calibration of predicted information gain. Predicted EIG is compared with observed information gain after action execution. The diagonal denotes perfect calibration. See also Figure S3.

Claim-level auditing showed that MedGenesis also produced fewer unsupported statements. Across evidence, reasoning, and action claims, its hallucination rates were 2.4%, 3.1%, and 0.8%, respectively. These rates were lower than Claude Code (6.2%, 7.4%, and 2.1%) and Codex CLI (7.8%, 9.4%, and 2.8%), and were far below the highest baseline error rates observed in Gemini 3 Pro, Grok 4.1, and Llama 4 (Figure 3C). Severity grading showed that most residual MedGenesis errors were harmless or minor, whereas several baseline systems accumulated a larger fraction of severe Score 2 errors. We then scored every hypothesis for falsifiability on a 0–5 rubric based on explicitly stated mechanism, direction of effect, measurable decision boundary, testable cohort or endpoint, and refutation condition. MedGenesis achieved the highest falsifiability distribution, with a median close to 4.5, compared with lower and broader distributions for all baselines (Figure 3D). Together, these results show that the Critic/Defender layer does not merely suppress unsupported claims; it also forces hypotheses into forms that can be tested.

We next asked whether MedGenesis could choose the next research action, not just write a better hypothesis. In the Latent Action Space, candidate actions were plotted by predicted EIG and UR. MedGenesis-selected actions clustered close to the feasible Pareto front and within the high-EIG/high-UR selected region, rather than collapsing into low-utility or infeasible areas (Figure 3A and Figure 3E). In the action-portfolio benchmark, each system ranked safe high-information actions, safe medium-information actions, low-value actions, generic underspecified actions, unsafe high-EIG near-misses, and abstention options. MedGenesis occupied the bottom-left optimal region of the ranking analysis, indicating low ranking error (1 − normalized discounted cumulative gain at 5 [NDCG@5]) and low top-1 regret (Figure 3F). This result means that MedGenesis placed high-utility actions near the top while avoiding unsafe or non-executable actions that only appeared informative. Finally, predicted EIG was prospectively calibrated against observed information gain after action execution. MedGenesis tracked the perfect-calibration diagonal most closely, whereas baseline systems systematically over- or under-estimated action value (Figure 3G). Figure 3 therefore links the benchmark advantage of MedGenesis to three measurable behaviors: it moves toward novel but plausible hypotheses, converts them into falsifiable claims, and selects safe actions whose predicted value matches realized information gain.

### MedGenesis generates clinical evidence from meta-analyses to case reports

We next deployed MedGenesis across five clinical evidence levels mapped onto the conventional clinical-evidence hierarchy (Figure 4A): systematic review and meta-analysis (L1), randomized trial subset analysis (L2), real-world data mining (L3), case-control study (L4), and case report (L5). Figure 4A also marks the wet-lab mechanistic branch as an L6 extension expanded in Figure 6. In each evidence-level task, the system received only a question and a dataset handle. Cohort assembly, analytical design, endpoint selection, and boundary reporting were generated autonomously and then reviewed post hoc by domain experts.

**Figure 4.**
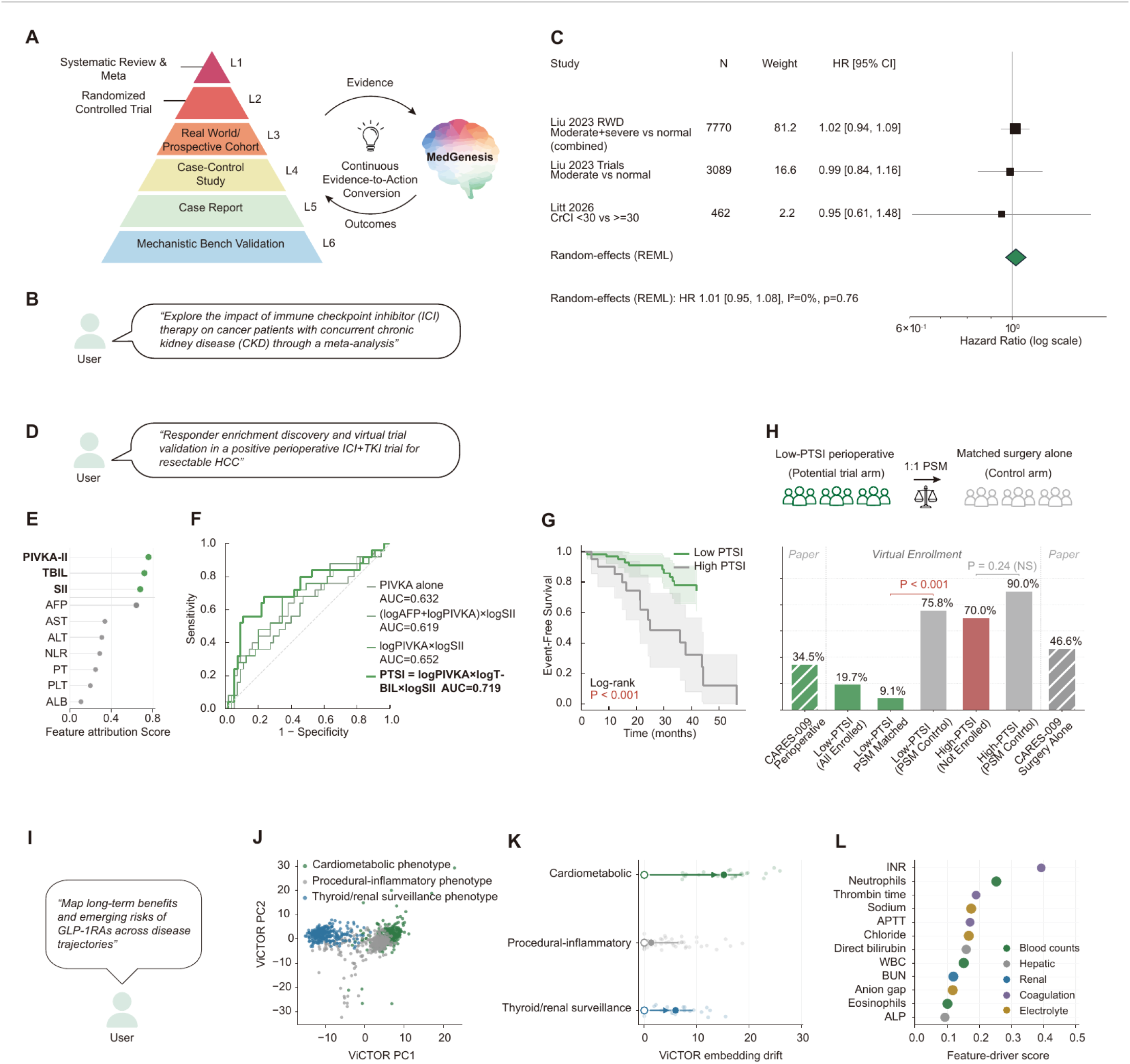
MedGenesis produces candidate clinical-research outputs across heterogeneous evidence formats. (A) Evidence hierarchy used to organize MedGenesis outputs. The figure spans systematic review and meta-analysis, randomized controlled trial analysis, real-world or prospective cohort analysis, case-control study, case report, and mechanistic bench validation. The MedGenesis loop links evidence to outcomes through continuous evidence-to-action conversion. (B) User prompt for the evidence-synthesis task, requesting a meta-analysis of immune checkpoint inhibitor (ICI) therapy in cancer patients with concurrent chronic kidney disease (CKD). (C) Main meta-analysis output for ICI therapy in CKD. The forest plot summarizes overall-survival hazard ratios from Liu 2023 real-world data, Liu 2023 pooled trials, and Litt 2026 severe renal-function subgroup data. (D) User prompt for the CARES-009 trial-enrichment task, requesting responder-enrichment discovery and virtual-trial validation in a positive perioperative ICI plus tyrosine kinase inhibitor trial for resectable hepatocellular carcinoma (HCC). (E) ViCTOR-assisted feature attribution for recurrence-associated pretreatment variables in the CARES-009 Zhongshan Hospital subset. PIVKA-II, total bilirubin (TBIL), systemic immune-inflammation index (SII), and alpha-fetoprotein (AFP) are shown with lower-ranked comparators. (F) Receiver operating characteristic comparison of recurrence biomarkers and formulas. PIVKA-II alone, (logAFP + logPIVKA-II) × logSII, logPIVKA-II × logSII, and the MedGenesis-derived pretreatment severity index (PTSI) were compared; PTSI achieved the highest AUC of 0.719. (G) Kaplan–Meier event-free survival analysis in the CARES-009 perioperative arm stratified by low versus high PTSI. The cutoff was selected by the Youden index. (H) Virtual-enrollment analysis based on PTSI. The event rate decreased from 34.5% in the unfiltered perioperative arm to 19.7% in low-PTSI patients and to 9.1% after propensity-score matching, compared with higher event rates in matched surgery-alone controls and high-PTSI patients. (I) User prompt for the real-world GLP-1 receptor agonist (GLP-1RA) trajectory task, requesting long-term benefit and risk mapping across disease trajectories. (J) ViCTOR patient-state map of 1,014 GLP-1RA-exposed patients at the index date. The map identifies cardiometabolic (n = 371), high-burden metabolic or procedural-inflammatory (n = 359), and renal/thyroid imaging-surveillance (n = 284) phenotypes. (K) ViCTOR embedding-drift distribution across the three GLP-1RA trajectory phenotypes between index and day-90 post-exposure states. Points show patient-level drift values within the cardiometabolic, procedural-inflammatory, and thyroid/renal surveillance phenotypes, with group-level summaries shown by colored markers. (L) Ranked patient-state-aware outcome signals among GLP-1RA-exposed patients. Decreased cardiometabolic events and increased imaging-detected thyroid or renal findings are shown as exploratory monitoring hypotheses rather than causal safety conclusions. Effect estimates are presented as hazard ratios with 95% confidence intervals (C), AUC values from ROC analysis (F), Kaplan–Meier estimates (G), event rates as percentages (H), and patient-level embedding-drift distributions (K). P values are displayed directly in the figure where shown, including non-significant results marked as NS. Statistical analyses include random-effects REML meta-analysis (C), ROC analysis (F), log-rank test (G), and chi-square or Fisher’s exact test after propensity-score matching (H), as appropriate. See also Figures S4 and S5.

We first asked whether chronic kidney disease (CKD) attenuates the survival benefit of immune checkpoint inhibitor (ICI) therapy in patients with cancer (Figure 4B). Because CKD reshapes antitumor immunity through uremic-toxin accumulation, chronic inflammation, impaired dendritic-cell function, and premature T-cell aging or exhaustion, its net effect on ICI benefit remains clinically uncertain^34,35^. We therefore asked MedGenesis to execute a PRISMA-style systematic review and meta-analysis of PubMed, Embase, and the Cochrane Library through 2024, including title-abstract screening, full-text review, and structured hazard-ratio extraction. The workflow screened 2,420 records, assessed 18 full-text reports, retained 15 studies for qualitative synthesis, and pooled three comparable overall-survival hazard ratio (HR) estimates^36,37^ (Figure S4A). A restricted maximum-likelihood (REML) random-effects model yielded a pooled HR of 1.01 (95% CI, 0.95–1.08; I^2^ = 0%; P = 0.76), indicating no overall attenuation of ICI benefit in patients with CKD (Figure 4C). Prespecified CKD-severity analyses showed pooled HRs of 0.94 (95% CI, 0.88–0.99) for mild CKD, 1.01 (95% CI, 0.94–1.08) for moderate CKD, and 1.12 (95% CI, 0.95–1.31) for severe CKD (Figure S4B). Because the severe-CKD estimate was driven by limited evidence, including one positive study by Tiu et al.^37^, MedGenesis flagged this subgroup as evidence-limited rather than ready for clinical extrapolation. The full run took 192 minutes and approximately 13 million tokens, converting a traditionally labor-intensive meta-analysis into an auditable, resource-tracked workflow.

We next asked whether MedGenesis could derive a pretreatment enrichment hypothesis for patients with resectable hepatocellular carcinoma (HCC) receiving perioperative camrelizumab, an anti–programmed cell death protein 1 antibody, plus rivoceranib, a vascular endothelial growth factor receptor 2 tyrosine kinase inhibitor, in CARES-009^38^ (Figure 4D). Before this trial, a phase 3 randomized clinical trial analysis of camrelizumab plus rivoceranib versus sorafenib in unresectable HCC had supported the clinical efficacy of this treatment backbone^39^. CARES-009 was the first positive randomized phase 2/3 perioperative immunotherapy trial reported in resectable HCC. In the intention-to-treat population, 294 patients were assigned to perioperative camrelizumab plus rivoceranib (n = 148) or surgery alone (n = 146), and perioperative treatment improved investigator-assessed event-free survival (EFS; HR, 0.59; 95% CI, 0.41–0.85; P = 0.0040)^38^. However, recurrence after perioperative therapy remained common, and no pretreatment rule was available to identify patients most likely to benefit. MedGenesis therefore used ViCTOR to compare pretreatment patient-state embeddings between recurrent and recurrence-free patients within the Zhongshan Hospital perioperative cohort (STAR Methods). Feature attribution prioritized protein induced by vitamin K absence or antagonist-II (PIVKA-II), total bilirubin (TBIL), systemic immune-inflammation index (SII), and alpha-fetoprotein (AFP). PIVKA-II, TBIL, and SII were integrated into the pretreatment severity index (PTSI) (Figure 4E; STAR Methods). PTSI showed the strongest recurrence stratification among tested candidates (Figure 4F and Figure S4C). PTSI was higher in recurrent patients (Mann–Whitney P = 0.002) and separated EFS at the Youden-optimal cutoff of 137, defining low PTSI as <137 and high PTSI as ≥137 (log-rank P < 0.001; Figure 4G and Figure S4D-E). In the virtual-enrollment analysis, selecting low-PTSI patients within the perioperative treatment arm reduced the EFS event rate from 34.5% in the unfiltered perioperative cohort to 19.7%. After 1:1 propensity-score matching with surgery-alone controls, the event rate further decreased to 9.1% in the matched low-PTSI perioperative subgroup, compared with 75.8% in matched surgery-alone controls (Figure 4H). Covariate balance after matching improved across age, sex, log(AFP + 1), log(PIVKA-II + 1), and log(SII + 1) (Figure S4F). These results support PTSI as a candidate pretreatment enrichment hypothesis and illustrate how MedGenesis turns patient-state geometry into a clinically interpretable trial-enrichment workflow, linking feature discovery, biomarker construction, survival stratification, matching, and virtual validation within a single auditable chain.

We then asked whether MedGenesis could use ViCTOR to transform glucagon-like peptide-1 receptor agonist (GLP-1RA) exposure from a medication label into interpretable patient-state trajectories (Figure 4I). Although GLP-1RAs have become highly effective therapies for metabolic disease and obesity, their long-term benefit-risk profile remains incompletely defined, particularly for cancer-related and thyroid/renal surveillance outcomes. Because recent real-world and multisite studies have reported mixed cancer-risk and thyroid-risk associations, MedGenesis treated these patterns as exploratory monitoring hypotheses rather than causal safety conclusions^40-42^. MedGenesis embedded index EHR snapshots from 1,014 GLP-1RA-exposed patients from Zhongshan Hospital and organized them into three clinically annotated phenotypes: Cardiometabolic (n = 371), procedural-inflammatory (n = 359), and thyroid/renal surveillance (n = 284) (Figure 4J). In 113 patients with paired 90-day post-exposure embeddings, ViCTOR quantified heterogeneous early state drift after treatment initiation, with 30, 61, and 22 patients from the three phenotypes, respectively (Figure 4K). MedGenesis then linked this drift to clinical features by integrating paired laboratory changes with broader EHR descriptors. The leading laboratory drivers spanned coagulation, inflammatory, electrolyte, hepatic, and renal domains, including international normalized ratio, neutrophils, thrombin time, sodium, activated partial thromboplastin time, chloride, direct bilirubin, white blood cells, blood urea nitrogen, anion gap, and alkaline phosphatase. The phenotype-explanation heatmap further showed cardiometabolic enrichment, procedural-inflammatory burden, and thyroid/renal surveillance intensity (Figure S4G). Thus, MedGenesis reframed GLP-1RA real-world analysis from a static exposure label into a dynamic, patient-state-aware map of clinical context and early treatment-associated movement.

We also used MedGenesis to perform a matched case-control analysis of severe immune-related adverse events (irAEs) in HCC patients receiving ICI therapy (Figure 5A). Because grade ≥3 irAEs can interrupt ICI therapy and are difficult to predict before treatment, MedGenesis was asked to derive a pretreatment monitoring hypothesis from routinely available baseline features. From 6,994 ICI-treated HCC patients in Zhongshan Hospital, 1,120 met the follow-up and complete-baseline-feature requirements. Among them, 101 developed grade ≥3 irAEs and were matched 1:4 to 404 controls. ViCTOR-assisted feature attribution prioritized portal vein tumor thrombus (PVTT), serum albumin, autoimmune history, and blood neutrophil-to-lymphocyte ratio (NLR) (Figure 5B). In the conditional logistic model, PVTT presence, autoimmune history, and elevated NLR were associated with increased severe-irAE risk, whereas higher albumin was protective (Figure 5C). In the simplified bedside score, NLR ≥3 and albumin <35 g/L were used as the prespecified clinical cutoffs. The model achieved an AUC of 0.715 and an optimism-corrected concordance index (C-index) of 0.693 (Figure 5D). MedGenesis converted these variables into an interpretable four-factor score using PVTT presence, albumin <35 g/L, autoimmune history, and NLR ≥3. The score stratified grade ≥3 irAE incidence from 9.9% to 15.4% and 35.5% across low-, intermediate-, and high-risk groups (Figure 5E). These findings turn baseline toxicity modeling into a bedside screening hypothesis for pre-ICI risk discussion and closer monitoring.

**Figure 5.**
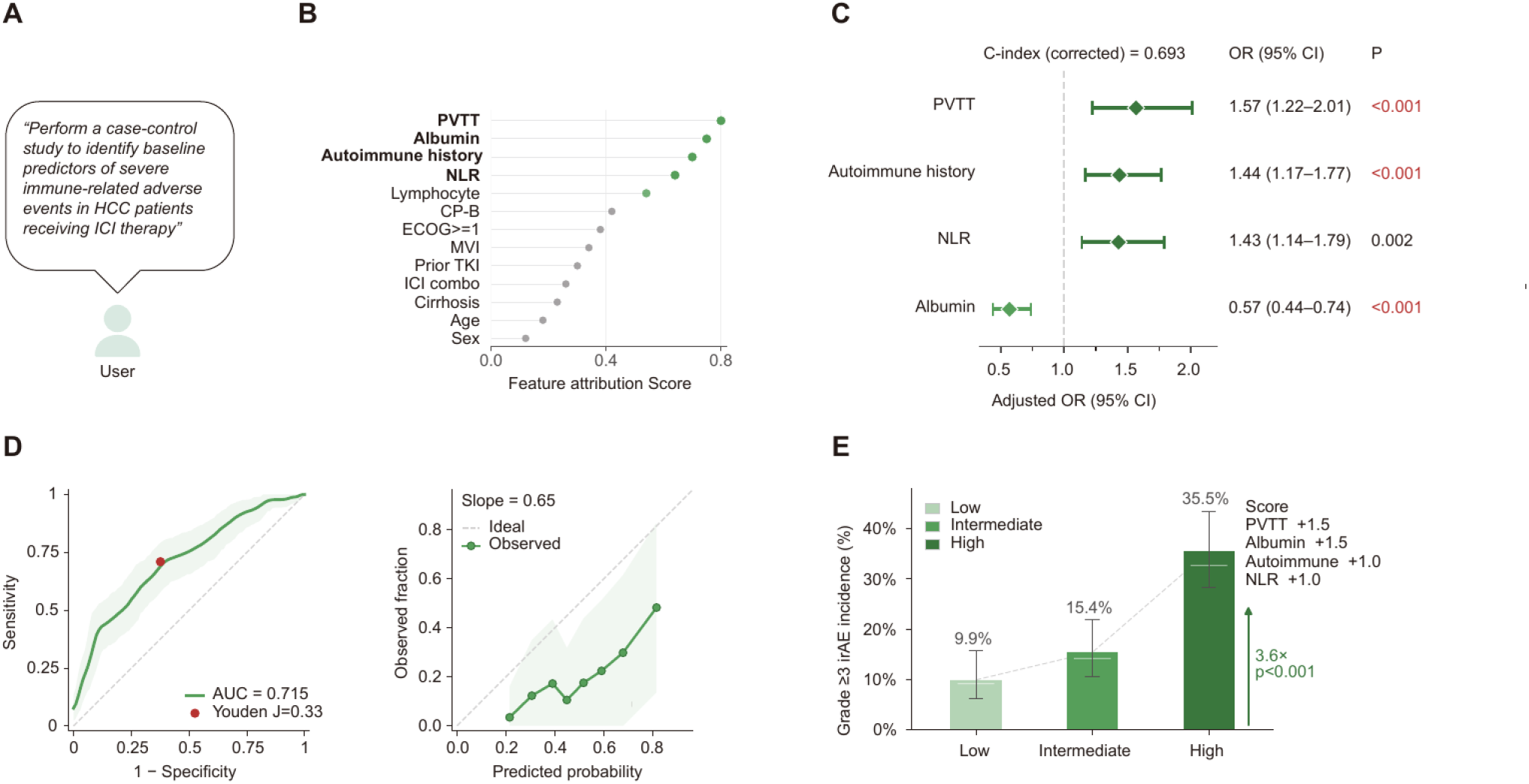
MedGenesis identifies baseline predictors of severe immune-related adverse events in HCC patients receiving immune checkpoint inhibitor therapy. (A) User prompt requesting a case-control study to identify baseline predictors of severe immune-related adverse events (irAEs) in HCC patients receiving immune checkpoint inhibitor (ICI) therapy. (B) ViCTOR-assisted feature attribution for severe irAE phenotyping. Portal vein tumor thrombus (PVTT), albumin, autoimmune history, and neutrophil-to-lymphocyte ratio (NLR) were prioritized for downstream model construction; additional clinical covariates are shown for comparison. (C) Adjusted-odds-ratio forest plot from the four-variable matched casecontrol model. PVTT, autoimmune history, and NLR increased severe-irAE risk, whereas albumin was protective. The optimism-corrected C-index was 0.693. (D) Discrimination and calibration of the irAE risk model. Left: receiver operating characteristic curve, AUC = 0.715, Youden J = 0.33. Right: observed versus predicted probability calibration plot, slope = 0.65. (E) Risk-stratified grade ≥3 irAE incidence under the weighted score. Low-, intermediate-, and high-risk groups had incidences of 9.9%, 15.4%, and 35.5%, respectively, corresponding to a 3.6-fold gradient. The simplified score assigned PVTT +1.5, albumin <35 g/L +1.5, autoimmune history +1.0, and NLR ≥3 +1.0. Effect estimates are presented as adjusted odds ratios with 95% confidence intervals (C), model performance as AUC, optimism-corrected C-index, Youden J, and calibration slope (D), and grade ≥3 irAE incidence as percentages with Wilson confidence intervals (E). P values are displayed directly in the figure where shown. Statistical analyses include conditional logistic regression (C), ROC and bootstrap calibration analyses (D), and chi-square trend test (E).

For rare-case discovery, MedGenesis used ViCTOR to screen 43,812 HCC patients from Zhongshan Hospital for comorbidity-associated cases (Figure S5A). A Wilson-disease-associated, viral-hepatitis-negative HCC case ranked first, supported by evidence for Wilson disease, portal hypertension, absence of viral hepatitis, and HCC (Figure S5B). MedGenesis reconstructed an 18-year timeline from Wilson disease diagnosis to portal-hypertension surgery, HCC diagnosis and treatment, tumor recurrence management, radiofrequency ablation, and transarterial chemoembolization (Figure S5C). A 12-marker immunohistochemistry panel supported hepatocellular differentiation and high proliferative activity, without overclaiming a copper-specific mutational mechanism (Figure S5D). This case was a rare but clinically coherent retrieval target: Wilson-disease-associated liver cancer is uncommon even in cirrhosis, with reported hepatobiliary malignancy prevalence of 1.2% and only two HCC cases among 130 patients followed for a median of 15 years^43,44^, whereas common cirrhosis etiologies carry annual HCC risks of approximately 2–3%^45^. Thus, MedGenesis converted large-scale chart search into an auditable case-report workflow that would otherwise require manual review of tens of thousands of records.

Together, these applications position MedGenesis as a clinical evidence translator: it converts fragmented institutional records into traceable claims about treatment benefit, monitoring hypotheses, toxicity risk, and rare-case value, each with an explicit boundary for prospective validation.

### Closed-loop refinement nominates a 3-hydroxybutyrate-neutrophil axis

To test whether MedGenesis could convert a broad biological question into an experimentally testable mechanism, we coupled it to a human-in-the-loop wet-lab workflow in tumor immunometabolism (Figure 6A). Beginning from the broad premise that tumor-associated metabolites may shape antitumor immunity, MedGenesis interrogated the UK Biobank metabolite panel and the published literature in parallel, scoring each of 249 circulating metabolites for epidemiologic association with incident cancer and mechanistic coverage in prior work. This prioritization was grounded in recent cancer-immunometabolism literature emphasizing metabolic control of immune state and treatment response^46^. The two streams converged on six ketone-related candidates. Among them, 3-hydroxybutyrate (3-HB) carried conflicting tumor-biology evidence: independent reports linked it to both tumor suppression and context-dependent tumor promotion (Figure 6B). MedGenesis flagged this contradiction as high EIG and prioritized 3-HB for follow-up. The accompanying epidemiologic association between circulating 3-HB and incident cancer was weak and non-linear, supporting controlled mechanistic testing rather than a direct population-level causal claim (Figure 6C).

**Figure 6.**
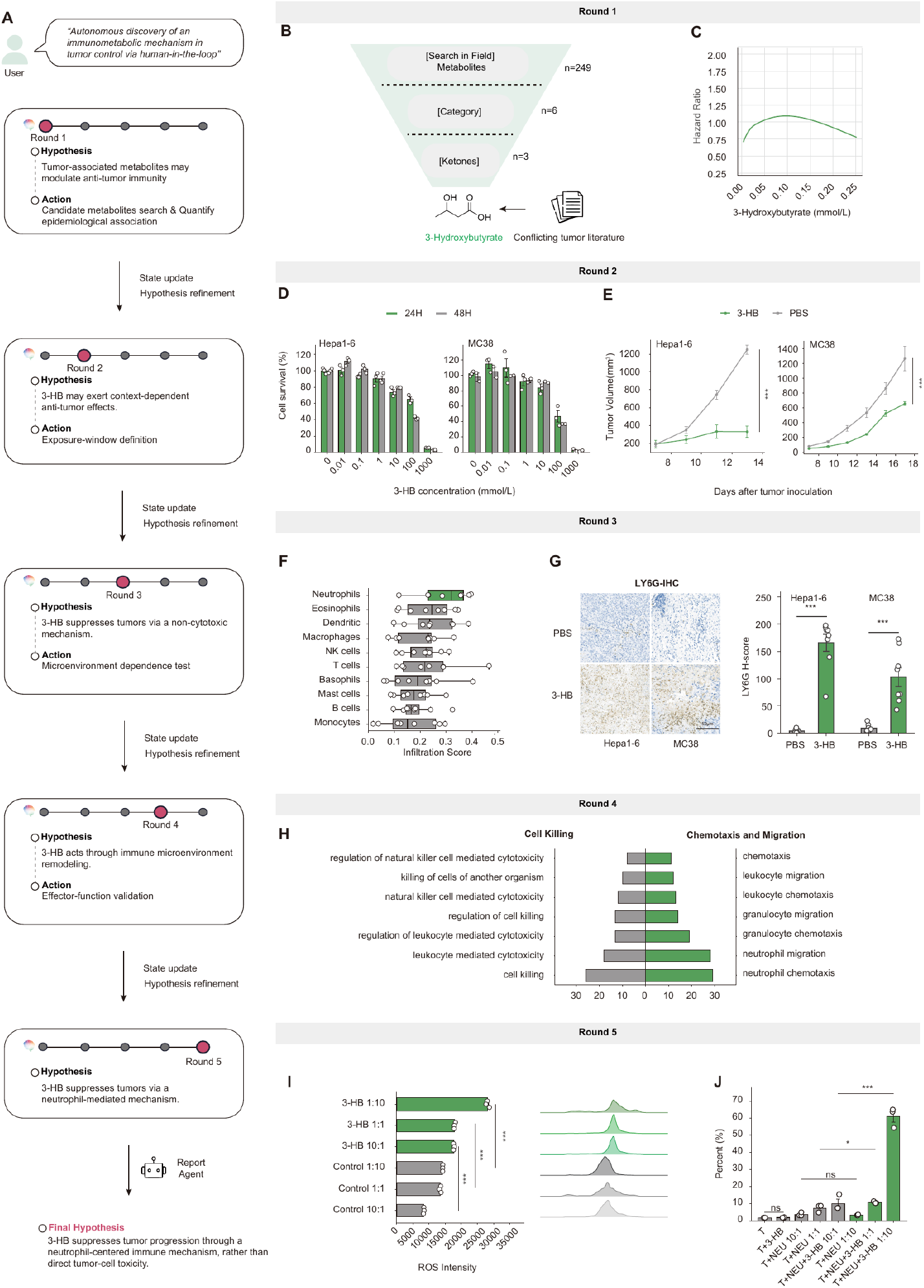
Closed-loop refinement nominates a 3-hydroxybutyrate-neutrophil tumor-suppression axis. (A) Human-in-the-loop mechanistic discovery trajectory. Across five refinement rounds, MedGenesis moved from a broad hypothesis that tumor-associated metabolites may modulate antitumor immunity to a final hypothesis that 3-hydroxybutyrate (3-HB) suppresses tumor progression through a neutrophil-centered immune mechanism rather than direct tumor-cell toxicity. Domain experts executed the selected animal and primary-cell experiments, and returned results were used for state updating. (B) Metabolite prioritization funnel. MedGenesis screened 249 circulating metabolites, narrowed the search to six ketone-related candidates and three ketone candidates, and prioritized 3-HB because of biological plausibility and conflicting tumor-literature evidence. (C) Epidemiologic association between circulating 3-HB exposure and incident cancer outcomes. The Cox regression hazard-ratio curve shows a weak, non-linear association across the observed 3-HB range, motivating controlled mechanistic testing. (D) Direct tumor-cell cytotoxicity testing. Hepa1-6 and MC38 cells were exposed to serial 3-HB concentrations for 24 h and 48 h. Cell survival was largely preserved at physiologic or ketogenic-range concentrations and decreased mainly at supraphysiologic concentrations. (E) In vivo tumor-growth experiment. C57BL/6 mice bearing syngeneic Hepa1-6 or MC38 tumors were treated with intratumoral 3-HB or PBS. 3-HB reduced tumor growth in both models. n = 10 mice per group. (F) Immune-infiltration prioritization. Pan-cancer immune-infiltration analysis using the Cancer Atlas of Metabolic Profiles and deconvolution consensus identified neutrophils as the highest-scoring 3-HB-associated immune population. (G) LY6G immunohistochemistry in PBS- and 3-HB-treated Hepa1-6 and MC38 tumors. Representative images and LY6G quantification show increased intratumoral neutrophil signal after 3-HB treatment. Scale bar, 50 µm. (H) Gene Ontology Biological Process enrichment in 3-HB-treated human peripheral-blood neutrophils compared with PBS-treated controls. Enriched programs include leukocyte-mediated cytotoxicity, regulation of cell killing, neutrophil migration, neutrophil chemotaxis, granulocyte migration, and chemotaxis. (I) Reactive oxygen species (ROS) intensity in neutrophil-tumor co-cultures with or without 3-HB across neutrophil-to-tumor ratios of 1:10, 1:1, and 10:1. (J) Propidium iodide (PI)-positive tumor-cell killing in the same co-culture system. Maximal tumor-cell death occurred in the combined Huh7 + neutrophils + 3-HB condition at a 10:1 neutrophil-to-tumor ratio, whereas Huh7 + 3-HB and Huh7 + neutrophils remained near background. Data in the bar plots are presented as mean ± standard deviation (D, G, I, and J), and tumor-growth curves are presented as mean ± standard error of the mean (E). Where shown in the figure, ns, not significant; *, P < 0.05; ***, P < 0.001. Statistical analyses include repeated-measures analysis or end-point Student’s t-test for tumor-growth curves (E), and Student’s t-test (G, I, and J). Transcriptomic enrichment statistics are reported as enrichment terms or normalized enrichment score/false discovery rate where applicable (H and Figure S6E). See also Figure S6.

Dose-response profiling was selected next as the action with the highest EIG. Across the murine discovery models Hepa1-6 and MC38 and the human HCC lines Huh7 and HCCLM3 (LM3), 3-HB had little or no effect at physiologic or keto-genic-range concentrations (≤10 mmol/L), with cytotoxicity emerging mainly at supraphysiologic concentrations (100– 1,000 mmol/L; Figure 6D; Figure S6B-C). When intratumoral 3-HB at 30 mmol/L was administered in syngeneic Hepa1-6 and MC38 tumors grown in immunocompetent C57BL/6 mice, tumor growth was significantly suppressed (P < 0.001; Figure 6E; Figure S6A). The dissociation between in vitro inertness and in vivo efficacy at the same concentration argued against direct tumor-cell cytotoxicity and led MedGenesis to redirect the next action toward the host immune microenvironment. Independent ketogenic-diet studies have linked ketone exposure to tumor-associated neutrophil polarization and immune-checkpoint response remodeling, consistent with an immune-mediated rather than purely cell-autonomous hypothesis^47^.

To identify the responsible immune compartment, MedGenesis interrogated the Cancer Atlas of Metabolic Profiles, comprising 988 specimens across 11 cancer types^48^, under a weighted consensus across seven immune-deconvolution methods. Neutrophils emerged as the lineage most consistently associated with 3-HB exposure (Figure 6F; Figure S6D). LY6G immunohistochemistry on Hepa1-6 and MC38 tumors confirmed a marked rise in intratumoral neutrophils after 3-HB treatment (Figure 6G), sharpening the working hypothesis from a general host-microenvironment effect to a neutrophil-centered mechanism. Recent reviews of neutrophil heterogeneity, neutrophil metabolism, and neutrophil-mediated drug resistance in cancer supported this lineage as a high-priority effector compartment^45-47^. MedGenesis next specified RNA sequencing (RNA-seq) of human peripheral-blood neutrophils treated with 3-HB or phosphate-buffered saline (PBS). Compared with PBS-treated neutrophils, 3-HB-treated neutrophils showed coordinated enrichment of chemotaxis, migration, leukocyte-mediated cytotoxicity, and cell-killing programs (Figure 6H; Figure S6E).

Closing the loop, MedGenesis specified a co-culture sufficiency test with four conditions: Huh7 cells alone, Huh7 + 3-HB, Huh7 + neutrophils, and Huh7 + neutrophils + 3-HB. These conditions were tested across neutrophil-to-tumor ratios of 1:10, 1:1, and 10:1. Reactive oxygen species (ROS) increased in neutrophil-containing cultures when 3-HB was present (Figure 6I). Propidium iodide (PI)-positive tumor-cell killing approached approximately 60% only in the combined Huh7 + neutrophils + 3-HB condition at the 10:1 ratio, where-as Huh7 + 3-HB and Huh7 + neutrophils remained near background (Figure 6J). These results support in vitro sufficiency of a 3-HB-conditioned neutrophil effector state, while leaving in vivo neutrophil-depletion and dose-bridging experiments for future validation. This mechanistic branch extends MedGenesis from clinical evidence generation to experimental mechanism discovery, linking a data-derived metabolite hypothesis to dose-response testing, syngeneic mouse models, immune-infiltration analysis, neutrophil transcriptomics, and co-culture sufficiency testing.

## DISCUSSION

This study presents MedGenesis as a clinical AI scientist that links hypothesis generation, action selection, skill execution, adversarial review, and memory update within one governed loop. Its main innovation is the explicit coupling of what is hypothesized with what is done next: candidate actions are ranked by EIG, UR, and P(safe), executed through auditable skills, and written back only after critic-defender review. Across clinical-research benchmarks, behavioral analyses, institutional patient datasets, and wet-lab-coupled validation, this design improved reasoning performance, reduced hallucination, and generated traceable candidate research products across heterogeneous evidence formats. By integrating ViCTOR as a patient-state modeling skill, MedGenesis also extends the loop from literature and structured skills to longitudinal patient trajectories, allowing EHR-derived cohorts, treatment histories, outcomes, and rare clinical patterns to enter the same hypothesis-action framework.

These findings should be viewed in the context of rapid progress in biomedical and clinical AI. Diagnostic systems such as DeepRare^4^ and MAI-DxO^5^ show that agentic reasoning can improve clinical diagnosis. Biological discovery systems such as OriGene^6^ and Biomni^7^ demonstrate that AI can coordinate target nomination, literature retrieval, and bioinformatics skill use. AI co-scientist frameworks show that multi-agent debate can improve hypothesis generation^8^. MedGenesis addresses a different unit of work: the end-to-end clinical research cycle. In clinical research, the difficult step is often deciding whether the next action should be a meta-analysis, cohort construction, subgroup enrichment, case-control modeling, trajectory analysis, rare-case retrieval, or wet-lab validation. This distinction is important for deployment because real clinical research systems must be extensible across skills, auditable across data sources, and conservative about claim boundaries. MedGenesis moves in this direction through the Skill Ocean, ViCTOR patient-state representations, and critic-defender governance, but it remains a research system. Its current outputs require expert review, external validation, prospective testing, and tighter integration with institutional data governance before clinical use.

The application studies illustrate the practical value of this framework. Across evidence synthesis, randomized-trial enrichment, real-world trajectory analysis, case-control risk scoring, and rare-case reporting, MedGenesis generated outputs with different statistical conventions rather than forcing every question into one template. The wet-lab-coupled 3-HB-neutrophil workflow further shows how the same loop can move from data-driven nomination to experimentally testable mechanism. Potential extensions include rare-disease diagnosis and case discovery, integration with imaging and pathology agents, automated multi-omics analysis, adaptive trial design, pharmacovigilance, and mechanism-guided patient stratification. The strongest near-term use case is therefore not autonomous clinical decision-making, but rapid generation of bounded, auditable research hypotheses that experts can prioritize for validation.

In summary, MedGenesis reframes clinical research from a sequence of disconnected expert workflows into a continuous evidence-to-action process. Its value lies in coupling what is hypothesized, what is done next, what evidence returns, and how claims are narrowed before they are reported. This provides a practical foundation for AI systems that assist clinical scientists not only by answering questions, but by helping decide which question, cohort, analysis, or experiment should come next.

### Limitations of the Study

This study has several limitations. MedGenesis was evaluated mainly in controlled benchmarks, retrospective cohorts, and research workflows, so its outputs require expert review, external validation, and prospective testing before clinical use. The CARES-009 PTSI, GLP-1RA trajectory signals, severe irAE risk score, and rare-case retrieval should be interpreted as candidate research hypotheses rather than validated clinical tools. The 3-HB-neutrophil mechanism was tested in selected models and co-culture systems, and further studies are needed to define dose, tumor-type specificity, immune dependency, and translational relevance.

## ACKNOWLEDGEMENTS

This study was supported by the National Natural Science Foundation of China (82130077, 82341008, 92459301, 82441049, 82394450), the Noncommunicable Chronic Diseases-National Science and Technology Major Project (2023ZD0502000), the Fundamental and Interdisciplinary Disciplines Breakthrough Plan of the Ministry of Education of China (No. JYB2025XDXM508), the Lingang Laboratory (LGL-8888-07), the Shanghai Academy of Natural Sciences (SANS) Exploration Scholars Project, and the New Cornerstone Science Foundation.

## AUTHOR CONTRIBUTIONS

Q.G., Y.C.W., P.T., S.J.Z., Z.F.Y., H.X., and N.J. conceived the MedGenesis framework and supervised the project. H.X., N.J., and T.C.Z. performed the literature review, designed the technical roadmap, constructed the benchmark strategy, and wrote the original draft. T.C.Z., J.C.G., Z.Y.Z., K.S., R.Y.W., Y.L., and Z.Z. contributed to the design of the AI foundation model architecture, ViCTOR patient-state modeling, benchmark construction, statistical workflows, figure generation, and data-governance SOPs. J.M.P. and J.Q.M. contributed to clinical cohort curation, endpoint adjudication, institutional data interpretation, and clinical validation. R.Y.W. and K.S. contributed to wet-lab experiments, animal studies, neutrophil functional assays, RNA-seq analyses, and mechanistic interpretation. All authors participated in the critical discussion, revision, and final approval of the manuscript. Q.G. served as the lead contact.

## STAR☆METHODS

Detailed methods are provided in the submitted files and include the following:

- **KEY RESOUCRCES TABLE**
- **RESOURCE AVAILABILITY**
  ○ Lead contact
  ○ Materials availability
  ○ Data and code availability
- **EXPERIMENTAL MODEL AND STUDY PARTICIPANT DETAILS**
  ○ Computational and clinical data resources
  ○ Human subjects and institutional clinical data
  ○ Controlled-access cohorts and EHR resources
  ○ Human donor neutrophils and generated RNA-seq samples
  ○ Cell lines
  ○ Mouse models
  ○ Ethics and governance
- **METHOD DETAILS**
  ○ MedGenesis architecture and closed-loop action selection
  ○ Composite action priority
  ○ Posterior update after action execution
  ○ Calibration of EIG and UR predictors
  ○ >Agent workflow and adversarial validation
  ○ Skill Ocean
  ○ ViCTOR patient-state representation model
  ○ ViCTOR evaluation, benchmark tasks, and attribution
  ○ ClinicalResBench and ClinicalRepBench construction
  ○ Benchmark evaluation and leakage control
  ○ Quantification of the Latent Hypothesis Space and Latent Action Space
  ○ Clinical hypothesis-generation benchmark
  ○ Novelty and plausibility scoring
  ○ Hallucination and severity scoring
  ○ Falsifiability scoring
  ○ Action selection and calibration benchmark
  ○ Action-space evaluation
  ○ Portfolio action-ranking evaluation
  ○ EIG calibration evaluation
  ○ Real-world clinical investigations
  ○ Meta-analysis of ICI therapy in cancer patients with CKD
  ○ CARES-009 PTSI pretreatment prognostic stratifier
  ○ GLP-1RA real-world disease-trajectory analysis
  ○ Severe immune-related adverse event feature discovery
  ○ Wilson-disease HCC rare-case retrieval and case construction
  ○ Wet-lab experimental validation workflow
  ○ Closed-loop wet-lab experimental design
  ○ Cell culture
  ○ Tumor-cell dose-response assays
  ○ Mouse tumor models and treatment
  ○ LY6G immunohistochemistry and immune-infiltration quantification
  ○ Human neutrophil isolation and 3-HB treatment
  ○ Human neutrophil RNA sequencing
  ○ Neutrophil RNA-seq processing and gene-set analysis
  ○ Neutrophil-tumor co-culture sufficiency assays
  ○ Flow cytometry
- **QUANTIFICATION AND STATISTICAL ANALYSIS**
  ○ Statistical analysis

## SUPPLEMENTAL INFORMATION

Supplemental information can be found in the submitted files.

## KEY RESOURCES TABLE

Key resources table is available in the submitted files.

## RESOURCE AVAILABILITY

### Lead contact

Further information and requests for resources, benchmark materials, code, and clinical workflow details should be directed to and will be fulfilled by the lead contact, Qiang Gao.

### Materials availability

This study did not generate new unique reagents outside the experimental materials described in the key resources table and Method details. Requests for study-specific prompts, benchmark construction templates, evaluation rubrics, and workflow configuration files should be directed to the lead contact.

### Data and code availability

#### Data availability

ClinicalResBench and ClinicalRepBench were constructed for this study. Benchmark metadata, task descriptions, scoring rubrics, aggregate model outputs, and processed evaluation tables will be made available upon publication through a public repository, subject to removal of copyrighted source text, restricted clinical content, and information that could enable re-identification of patients or reconstruction of institutional records. Full paper-reproduction task materials in ClinicalRepBench will be released in redacted form when redistribution of original article content is not permitted.

Publicly available datasets and resources used in this study can be accessed from their original repositories, including PubMed/PMC, TCGA, GEO, GTEx, CPTAC, SEER, NHANES, ClinicalTrials.gov, ChEMBL, PubChem, FAERS, and other cited public databases. Controlled-access resources, including UK Biobank and MIMIC-IV, remain governed by their original data-use agreements and must be accessed through their respective application procedures. The UK Biobank analysis was conducted under an approved UK Biobank access application.

Institutional EHR, pathology, imaging, follow-up, and outcome-registry data from Zhongshan Hospital, Fudan University, are not publicly available because they contain patient-level clinical information and are subject to institutional governance, ethics approval, and data-use restrictions. All institutional analyses were performed within the approved institutional computing environment. No patient-level institutional data left the institutional infrastructure. De-identified aggregate results, summary statistics, model-evaluation tables, and figure-level processed data will be made available where permitted by ethics approval and data-governance policy.

#### Code availability

Analysis code used to generate the figures, benchmark summaries, statistical analyses, meta-analysis outputs, survival analyses, enrichment analyses, and processed tables will be released in a public repository upon publication. The repository will include documented scripts, environment files, example inputs, and instructions sufficient to reproduce the reported aggregate analyses from released or publicly obtainable data.

Code for ClinicalResBench and ClinicalRepBench evaluation, including scoring templates, rubric files, post-processing scripts, and aggregate benchmarking notebooks, will be released after removal of restricted source text and copyrighted or non-redistributable materials. Code for analyses involving controlled-access cohorts or institutional EHR data will be provided as executable templates or pseudonymized workflow scripts, but not with patient-level inputs. Components that depend on protected institutional databases, governed EHR wrappers, or internal access-control systems will not be publicly released.

## EXPERIMENTAL MODEL AND STUDY PARTICIPANT DETAILS

### Computational and clinical data resources

Computational and clinical data resources included institutional human-subject records, controlled-access cohorts, public clinical resources, pharmacovigilance resources, and public multi-omics datasets. These resources supported MedGenesis agent execution, ViCTOR patient-state modeling, benchmark construction, real-world cohort analyses, UK Biobank metabolite prioritization, and public tumor-context analyses.

### Human subjects and institutional clinical data

Institutional human-subject data came from Zhongshan Hospital, Fudan University, and were used only after de-identification under local governance for secondary research. The institutional data layer included structured EHRs, diagnoses, laboratory measurements, medication orders, procedures, treatment records, imaging and pathology reports, adverseevent records, follow-up registries, and outcome registries. These resources supported ViCTOR patient-state modeling, clinical cohort retrieval, trajectory stratification, risk-factor discovery, rare-case retrieval, and endpoint analyses.

The ViCTOR patient-state corpus contained 103,812 institutional patients. The CARES-009 analysis used the Zhongshan Hospital subset of the multicenter perioperative HCC trial (NCT04521153), including 106 patients in the perioperative treatment arm and 124 patients in the surgery-alone arm. The GLP-1RA trajectory analysis included 1,014 exposed patients with eligible longitudinal EHR follow-up. The severe irAE analysis screened 6,994 ICI-treated HCC patients, retained 1,120 eligible patients after follow-up and baseline-feature completeness criteria, and analyzed 101 severe-irAE cases matched to 404 controls. The Wilson-disease HCC retrieval task searched an institutional embedding index containing 43,812 HCC patients.

### Controlled-access cohorts and EHR resources

Controlled-access resources were handled as study-participant or EHR resources according to their native data-use agreements. UK Biobank metabolite and phenotype data were used for the circulating-metabolite prioritization step in the wet-lab-coupled discovery branch. MIMIC-IV was used as a controlled-access EHR resource available to the MedGenesis data layer and benchmark-oriented clinical reasoning workflows. These controlled resources were not redistributed; analyses were performed through approved access routes, and only aggregate or derived results were retained in the manuscript output.

### Human donor neutrophils and generated RNA-seq samples

This study generated a human neutrophil RNA-seq dataset for the 3-HB mechanistic branch. Fresh peripheral-blood neutrophils were isolated from de-identified healthy donors and assigned to PBS vehicle or 3-HB treatment ex vivo. The experimental comparison was prespecified as 3-HB-treated neutrophils versus PBS-treated neutrophils. Donor and replicate identifiers, sample-processing records, RNA-quality records, and processed expression matrices are retained in the Bio-Med Big Data Center (https://www.biosino.org/node/home).

### Cell lines

Murine Hepa1-6 hepatoma cells and MC38 colon carcinoma cells were used for syngeneic mouse tumor experiments and in vitro dose-response testing. Huh7 and HCCLM3 (LM3) human hepatocellular carcinoma cells were used for dose-response and neutrophil-tumor co-culture experiments. Cells were maintained under standard mammalian culture conditions at 37°C in a humidified incubator with 5% CO2 and were confirmed to be mycoplasma-free before animal implantation or functional assays. Cell-line authentication and culture records are retained in laboratory documentation.

### Mouse models

C57BL/6 mice were used for immunocompetent syngeneic tumor models. Hepa1-6 and MC38 cells were implanted subcutaneously, and mice were assigned to PBS vehicle or 3-HB treatment after tumors became measurable. For each tumor model, 10 mice were assigned to PBS and 10 mice were assigned to 3-HB treatment. Tumors were collected for growth assessment, histology, LY6G immunohistochemistry, and immune-infiltration analysis. Mice were housed under specific pathogen-free conditions with controlled temperature, humidity, and light-dark cycle, with free access to food and water.

### Ethics and governance

The study was approved by the Institutional Review Board of Zhongshan Hospital, Fudan University (No. B2023-350). Human analyses were conducted in accordance with the Declaration of Helsinki and institutional data-governance rules. De-identified donor blood samples were handled under the same approved governance framework. Animal experiments were performed under institutional animal-care procedures, with humane endpoints applied according to approved laboratory practice.

## METHOD DETAILS

### MedGenesis architecture and closed-loop action selection

MedGenesis was designed as a closed-loop clinical discovery system rather than a single-pass question-answering model. At each iteration, the system maintains a current hypothesis set, a clinical world state, and a set of candidate actions that can be executed through governed skills, including literature retrieval, cohort construction, survival analysis, biomarker discovery, causal/statistical modeling, trial emulation, patient-state modeling, and wet-lab handoff. A candidate action may be a database query, a statistical analysis, a model call, a literature synthesis step, a case-retrieval step, or a proposed validation experiment.

### Composite action priority

Action selection was governed by a Clinical World Model. For each candidate action a, MedGenesis estimates a composite priority score:

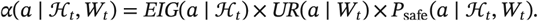

where EIG and UR denote the previously defined action-priority terms, and P(safe) is a governance prior that downweights actions violating clinical, ethical, privacy, or feasibility constraints. EIG was defined as:

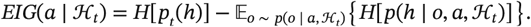

where h indexes competing hypotheses and o denotes the possible observation returned by action a. The entropy term was computed as:

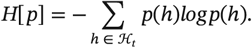

UR was estimated as:

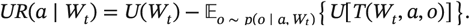

### Posterior update after action execution

where U(Wt) is the uncertainty of the current world state and T(Wt, a, o) is the state transition after observing o. After execution, the posterior belief over hypotheses was updated by:

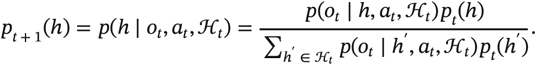

### Calibration of EIG and UR predictors

Predicted EIG and UR were calibrated against observed changes in hypothesis entropy and state uncertainty across completed actions. The calibration loss was:

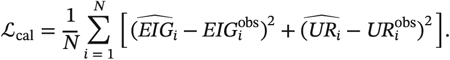

This formulation allowed MedGenesis to prioritize actions that were not only plausible but discriminative, executable, and clinically governed.

### Agent workflow and adversarial validation

The system was implemented as a coordinated multi-agent workflow. A Scout Agent defines the discovery space, a Hypothesis Agent converts open-ended clinical questions into structured and falsifiable hypotheses, an Evidence Agent assembles the current data state, an Action Agent ranks candidate next steps under the composite action score, and an Execution Agent calls the selected skills. The resulting evidence is then passed to a Critic-Defender adversarial validation layer, where claims are tested for confounding, selection bias, leakage, overinterpretation, internal contradiction, and unsupported causal language. A Report Agent writes the accepted evidence trace, failed alternatives, and residual uncertainty back to long-term memory before the next iteration.

The multi-agent layer was implemented using LangGraph and the Claude Agent SDK. LangGraph defined the stateful orchestration graph, including agent nodes, conditional routing, checkpointing, and handoff across hypothesis generation, evidence integration, action selection, execution, adversarial validation, and report writing. The Claude Agent SDK provided the agent runtime for instruction following, structured tool invocation, schema-constrained output parsing, and inter-agent communication. Each agent was configured with a role-specific instruction, permitted tool set, input-output schema, and governance constraints, and all tool calls, intermediate states, Critic-Defender exchanges, and memory-write decisions were recorded in auditable execution logs. Patient-level institutional data remained inside the approved local computing environment.

### Skill Ocean

The Skill Ocean comprised 858 callable skills organized into six functional families aligned with clinical-scientist capabilities rather than software packages. These included literature retrieval and evidence synthesis (143 skills; PubMed/PMC retrieval, citation-graph traversal, cross-database search, evidence extraction, guideline retrieval), clinical population and recruitment modeling (139 skills; EHR cohort building, ClinicalTrials.gov search, ViCTOR embedding, patient-state stratification, virtual recruitment), molecular and omics follow-up (147 skills; gene/protein lookup, pathway analysis, structure and variant search, TCGA/GTEx/GEO query, biomarker discovery), drug, compound and safety intelligence (143 skills; Food and Drug Administration (FDA) drug-label mining, absorption, distribution, metabolism, excretion, and toxicity (ADMET) prediction, ChEMBL/PubChem search, drug-target interaction analysis, FDA Adverse Event Reporting System (FAERS) safety-signal detection), modeling, embedding and analytics (143 skills; statistical execution, causal inference, model-embedding profiling, feature-finding workflows, target-trial emulation), and study design and evidence governance (143 skills; endpoint definition, claim governance, trial-design simulation, evidence-trace assembly, hallucination audit). Each skill exposed a standardized input-output schema, execution log, provenance record and failure mode, allowing MedGenesis to assemble the task-specific world state on demand and to route every executed action through traceable evidence and governance checks. This structure matches the updated Figure S1A Skill Ocean panel, which lists the six families and their skill counts.

### ViCTOR patient-state representation model

ViCTOR was used as the patient-state representation layer of MedGenesis, converting irregular institutional EHRs into longitudinal embeddings that could be queried by downstream clinical-research workflows. The model was designed for settings in which diagnoses, laboratory measurements, treatments, procedures, imaging reports, pathology descriptions, and follow-up events occur at uneven intervals and cannot be reduced reliably to a single fixed feature table. For each patient, ViCTOR constructed time-ordered clinical-state windows and produced embeddings at prespecified index dates. MedGenesis then used these embeddings for cohort comparison, patient matching, trajectory retrieval, risk attribution, and downstream prediction.

The ViCTOR training corpus comprised approximately 558,683 clinical visits, 545,896 longitudinal patient-state windows, and 185.3 million event-value-time tokens. Patient-level splitting was used to avoid leakage between training, validation, and test sets, yielding approximately 439,179 training windows, 54,647 validation windows, and 52,070 test windows. Each patient-state window contained a mean of 278.6 events and a median of 316 events, providing longitudinal context for both near-term clinical states and longer disease trajectories.

Structured inputs included diagnosis events, laboratory measurements, examinations, medications, surgeries and procedures, report-derived clinical events, and special temporal or structural tokens. At the 103,812-patient scale, the event corpus included approximately 2.94 million diagnosis tokens, 682,282 examination tokens, 48.99 million laboratory tokens, 3.58 million medication tokens, 839,370 surgery tokens, 9.06 million text-derived event tokens, and 1.03 million special tokens. Numeric laboratory values were unit-harmonized when possible, transformed or binned to reduce skewness, and encoded with their source event metadata. Categorical variables were canonicalized before tokenization.

Narrative clinical information was incorporated as embedded observations rather than as raw text. Clinical notes, imaging reports, and pathology reports were segmented into report-level text chunks, encoded with PubMedBERT, and projected into the same patient-state space as structured events. This text module contained approximately 226,128 visit-level text embeddings derived from 226,186 text chunks, each represented initially as a 768-dimensional PubMedBERT embedding. The resulting representation allowed ViCTOR to include narrative findings while preserving a uniform longitudinal event sequence.

ViCTOR used a Transformer-based encoder-decoder architecture. Each structured event was represented by an event type, clinical code, value or value bin, and temporal position. Embedded reports and other unstructured observations were passed through modality-specific projection layers and combined with learnable time encodings. The ordered event sequence was processed by the Transformer encoder to produce contextual patient-state vectors, and patient-level embeddings were extracted at prespecified index dates from the final clinical state or task-specific query representation.

Pretraining used masked event modeling and temporal reconstruction objectives. Randomly selected structured codes, value bins, and report embeddings were masked, and the decoder was trained to reconstruct the missing information from surrounding longitudinal context. Structured codes used classification losses, numeric or binned values used regression or classification losses as appropriate, and report embeddings used distance-based reconstruction losses. Auxiliary time-to-event objectives were included when endpoint labels were available. Model selection used validation loss and downstream validation performance, and final evaluations were performed only on held-out patients.

### ViCTOR evaluation, benchmark tasks, and attribution

ViCTOR was evaluated to test whether its embeddings captured clinically meaningful patient states rather than only local visit information. These 226 tasks comprised 109 new-onset disease prediction tasks, 63 disease-progression prediction tasks, and 54 treatment-efficacy prediction tasks across seven retained disease families: cardiovascular and circulatory, endocrine and reproductive, gastrointestinal/hepatic/renal, hematology and sensory, musculoskeletal and autoimmune, respiratory and infectious, and oncology. Frozen ViCTOR embeddings were paired with lightweight task-specific prediction heads and compared with age-sex reference models. Discrimination was summarized with area under the receiver operating-characteristic curve (AUROC), area under the precision-recall curve (AUPRC), and C-index according to end-point type.

A second benchmark assessed patient-level retrieval. Candidate diagnosis-medication and disease-trajectory phenotypes were first generated from the institutional EHR, then filtered for adequate positive query counts and clinical interpretability. The final manuscript retrieval panel retained 45 selected tasks spanning six disease families: HCC and metastatic cancer, cardiovascular and circulatory disease, endocrine/metabolic disease, respiratory and infectious disease, hepatic/gastrointestinal/renal disease, and hematology/bleeding disorders. For each task, positive test patients served as queries and were removed from the candidate pool before ranking. Candidate patients were ranked by embedding similarity, and retrieval performance was compared with a latest-progress-note retrieval baseline using accuracy at 5 (Acc@5), with mean average precision reported where applicable.

For interpretability, ViCTOR attribution was calculated at the event and feature-family levels. For each downstream task, the encoder was frozen and a lightweight task head was fitted on extracted patient-state embeddings. Attribution from the task output back to the longitudinal input sequence was estimated using integrated-gradient or perturbation-based embedding attribution, then mapped back to clinically interpretable variables including diagnoses, laboratory tests, medications, procedures, report-derived features, and temporal visit patterns. Scores were normalized within each task so that displayed values reflected relative feature contribution rather than absolute clinical effect size.

MedGenesis used this attribution layer to guide feature discovery and hypothesis checking in the CARES-009 recurrence analysis, severe irAE risk modeling, GLP-1RA trajectory analysis, and Wilson-disease HCC case retrieval. In each application, ViCTOR supplied patient-state representations and interpretable evidence, but the final clinical claim required downstream statistical testing, governance review, and domain-expert interpretation.

### ClinicalResBench and ClinicalRepBench construction

ClinicalResBench was constructed to evaluate clinical-research reasoning beyond conventional medical question answering. The benchmark contains 1,697 expert-curated question-answer pairs organized into two complementary tracks. ClinicalRes-Knowledge contains 1,256 questions covering 10 clinical domains, including oncology, cardiology, neurology, infectious diseases, endocrinology, immunology, respiratory medicine, gastroenterology, hematology, and dermatology. ClinicalRes-Methodology contains 441 questions covering 5 study-design categories, including randomized controlled trials (RCTs), cohort studies, systematic reviews and meta-analyses, case-control studies, and case series.

Candidate questions were generated from clinical literature, practice guidelines, translational-medicine reports, real-world evidence studies, and representative clinical-research scenarios. The construction workflow included source selection, LLM-assisted candidate drafting, de-duplication, ambiguity filtering, non-triviality screening, answer verification, and final expert adjudication. Questions answerable by simple lexical retrieval, questions with multiple defensible answers, questions depending on hidden assumptions, and questions with insufficient evidence support were removed or revised. Final answers were paired with structured scoring rubrics specifying required concepts, acceptable alternatives, common errors, and citation requirements.

ClinicalRepBench was constructed to evaluate whether an AI system could reproduce and extend published clinical-research workflows rather than only answer isolated questions. The benchmark includes 40 paper-reproduction tasks across 10 clinical domains, with 4 representative studies selected per domain. Each task required the system to identify the clinical question, reconstruct the study design, specify the analytic population and endpoints, reproduce the key statistical or evidentiary reasoning, and generate an evidence-traced interpretation. Tasks were designed to include both Re-Discovery and New-Discovery components: Re-Discovery assessed whether the system could recover the main reported finding of the source study, whereas New-Discovery assessed whether it could identify a plausible additional signal, subgroup, or limitation from the same evidence space without overclaiming.

ClinicalRepBench outputs were scored on a 0–100 scale across six dimensions: quantitative reasoning, statistical understanding, clinical-domain knowledge, evidence synthesis, critical appraisal, and causal reasoning. A score of 50 was defined as the approximate human-expert reference level for a complete but not exceptional reproduction. Scores below this threshold reflected incomplete reconstruction, missing assumptions, weak statistical reasoning, or insufficient evidence traceability. Scores above this threshold required accurate reconstruction plus clinically meaningful critique, appropriate uncertainty boundaries, and justified extension beyond the reported result. Final task scores were averaged within domains and then across domains to obtain the overall ClinicalRepBench score.

### Benchmark evaluation and leakage control

MedGenesis was evaluated against three classes of baselines under matched prompting and execution settings: frontier general-purpose LLMs, an existing biomedical multi-agent system, and code-executing agents. For ClinicalResBench, each system received the same question text and was asked to provide a direct answer with supporting reasoning and evidence trace when applicable. For ClinicalRepBench, each system received the same paper-reproduction task description and was asked to reconstruct the study logic, analytic workflow, key results, interpretation, and limitations. Code-executing agents were allowed to use executable analysis workflows when the task required computation, whereas non-code systems were evaluated on their reasoning and reconstruction outputs.

ClinicalResBench answers were scored using the expert-approved rubrics developed during benchmark construction. ClinicalRepBench outputs were scored using paper-specific reproduction rubrics and the six-dimensional evaluation framework described above. Free-text outputs were reviewed by blinded adjudicators who were not shown the model identity during scoring. Borderline or discordant cases were re-reviewed by an additional domain expert, and final scores were assigned after consensus adjudication. Systems were penalized for unsupported factual claims, fabricated citations, numerical errors, missing provenance, causal overclaiming, inappropriate extrapolation, failure to identify key assumptions, and failure to distinguish exploratory findings from confirmatory evidence.

To evaluate test-time scaling, systems were run under increasing compute budgets. For MedGenesis, additional compute was allocated to additional hypothesis–action refinement rounds, deeper evidence retrieval, expanded adversarial validation, and repeated evidence-trace checking. For baseline LLMs, additional compute was implemented through matched repeated sampling, self-consistency, or extended reasoning where supported. For code-executing agents, additional compute was allocated to longer execution time, iterative code revision, and repeated analysis attempts. Wall-clock time, approximate compute budget, and final task score were recorded to compare accuracy–time–compute trade-offs across systems.

Leakage control was performed at both the question-answer and paper-reproduction levels. For ClinicalResBench, candidate items were screened for verbatim overlap between the question, answer, and widely indexed source text. Items answerable by direct title matching, abstract memorization, or isolated keyword retrieval were removed or rewritten. For ClinicalRepBench, tasks were constructed to require reconstruction of study design, analytic logic, and uncertainty boundaries rather than simple recall of a paper conclusion. Source papers were checked for trivial answer leakage, and prompts did not include the exact paper title when doing so would make retrieval alone sufficient. Where a source study was publicly available, scoring emphasized reproducible reasoning, methodological reconstruction, and critical appraisal rather than whether the model could recall the reported conclusion.

Additional safeguards were applied to reduce evaluation bias. All systems were evaluated with standardized instructions, matched output requirements, and comparable opportunities to provide evidence. Sampling parameters were harmonized within model families when configurable. The benchmark was locked before final model runs, and no scored outputs were used to revise prompts, rubrics, or task definitions. Because both ClinicalResBench and ClinicalRepBench were constructed for this study, they should be interpreted as study-specific research benchmarks rather than externally established community benchmarks; however, their expert-curated design, leakage screening, blinded adjudication, and cross-domain composition were intended to provide a controlled and clinically realistic evaluation of end-to-end clinical-research competence.

### Quantification of the Latent Hypothesis Space and Latent Action Space

#### Clinical hypothesis-generation benchmark

We constructed a clinical hypothesis-generation benchmark to evaluate whether MedGenesis and comparator systems could produce hypotheses that were novel, clinically plausible, evidence-grounded, and experimentally refutable. The benchmark included 18 clinical-research tasks spanning oncology, metabolic medicine, cardiology, neurology, critical care, obstetrics, gastroenterology/immunology, transplant medicine, infectious disease, geriatrics, rheumatology/nephrology, pediatrics, rare disease genetics, and psychiatry/metabolic medicine. Each task contained a clinical question, a short evidence packet defining allowable task-specific facts, and an unverified trap statement designed to test whether a model would repeat unsupported clinical claims.

The evaluated systems were MedGenesis, Claude Code, Codex CLI, Biomni, Gemini 3 Pro, GPT-5.2, Claude 4.5 Opus, Grok 4.1, DeepSeek-V3.2-Thinking, and Llama 4. API-accessible baseline models were queried through OpenRouter using the same task prompts. Each model generated one output per task, yielding 18 outputs per model. Outputs were anonymized before scoring so that judge models and human adjudicators evaluated only the task context and candidate text.

#### Novelty and plausibility scoring

For Figure 3B, each output was scored for novelty and plausibility on a 0-1 scale. Novelty was defined as clinically meaningful non-obviousness rather than unsupported creativity. Plausibility was defined as biological and clinical credibility under the evidence packet. To reduce sensitivity to style, verbosity, and model identity, outputs within each task were also ranked in blinded candidate comparisons.

The final novelty and plausibility values combined absolute judge scores with blinded ranking scores. Penalties were applied for hallucinated evidence, unsupported causal reasoning, unsafe or underspecified actions, and low falsifiability. For each model, scores were summarized across the 18 tasks as the median and interquartile range. In Figure 3B, open circles indicate novelty and filled circles indicate plausibility. Scores of at least 0.75 were treated as the predefined Ideal zone.

#### Hallucination and severity scoring

For Figure 3C, judges audited each model output at the claim level. Claims were grouped into three categories: Evidence claims, Reasoning claims, and Action claims. Evidence claims captured factual support, citations, study results, sample sizes, or trial identifiers. Reasoning claims captured causal or mechanistic interpretation. Action claims captured the feasibility, safety, and executable structure of the proposed clinical-research action.

Each representative claim was assigned a severity score of 0, 1, or 2. Score 0 indicated harmless or supported content. Score 1 indicated minor unsupported or weakly overclaimed content. Score 2 indicated severe fabricated, unsafe, infeasible, or clinically misleading content. Category-specific hallucination rates were calculated as the percentage of audited claims with severity greater than 0 within each category. Severity distribution was calculated per output using the maximum severity observed across Evidence, Reasoning, and Action claims, then summarized as the percentage of outputs with Score 0, Score 1, or Score 2 for each model.

#### Falsifiability scoring

For Figure 3D, falsifiability was scored on a 0-5 rubric based only on explicitly stated design elements. One point was awarded for each of the following: a specified mechanism or rationale, a specified direction of effect, a measurable threshold or operational decision boundary, a testable cohort or endpoint, and an explicit refutation or failure condition. Judges were instructed not to infer missing details from conventional study design.

The distribution of falsifiability scores across the 18 tasks was plotted for each model as a violin plot with sample-level points and median/interquartile-range overlays. Higher scores indicated that a hypothesis could be translated directly into a cohort analysis, trial-emulation design, meta-analysis, mechanistic experiment, or validation study.

#### Action selection and calibration benchmark

For Figure 3E-G, we evaluated whether MedGenesis could convert candidate biomedical hypotheses into executable, safe, informative, and well-calibrated research actions. Each task contained a biomedical research question, competing hypotheses, a task-specific evidence bundle, and unresolved uncertainty components. The evaluation focused on the hypothesis-to-action step rather than final-answer correctness. The benchmark therefore measured whether a model selected actions that were specific, testable, safe, hypothesis-discriminating, and prospectively calibrated.

For each task, MedGenesis generated a latent action space. Each candidate action was represented by an action type, natural-language description, structured parameters, predicted EIG, predicted UR, and P(safe). Baseline systems were evaluated on the same candidate action portfolios and ranked candidate actions for execution. Actions were down-weighted or blocked when they had patient-level privacy risk, proposed non-permitted data export, implied prospective clinical intervention from an exploratory signal, lacked endpoint or comparator definition, omitted a time window, or failed to specify adequate confounder or governance control.

#### Action-space evaluation

Figure 3E evaluated whether MedGenesis selected actions from the high-value region of the action space. Candidate actions were plotted by predicted EIG and predicted UR. The ideal region corresponded to actions with both high expected information gain and high uncertainty reduction. Low-utility actions had low EIG and low UR. Actions with high apparent EIG but poor feasibility, safety, or executable structure were not considered optimal.

The reference Pareto front was computed from feasible candidate actions after excluding blocked or unsafe actions. An action was considered Pareto optimal if no other feasible action had both higher EIG and higher UR. The MedGenesis-selected action was then compared with this feasible Pareto front. A good selector was expected to choose actions close to the Pareto front and inside the high-EIG/high-UR region, rather than selecting generic, low-yield, unsafe, or non-executable actions.

#### Portfolio action-ranking evaluation

Figure 3F evaluated whether each model could rank candidate research actions in the correct execution order. For each task, we constructed an action portfolio containing safe high-information actions, safe medium-information actions, low-value safe actions, generic underspecified actions, unsafe high-EIG near-miss actions, and abstention actions. The candidate portfolios were designed to distinguish models that simply favored high nominal EIG from models that recognized actions that were safe, executable, and likely to produce useful evidence.

Each action was scored after execution using observed information gain, observed uncertainty reduction, execution yield, result success, result richness, parameter specificity, governance compliance, and bias-control quality. Ranking quality was measured using normalized discounted cumulative gain at 5 and top-1 regret. In Figure 3F, ranking error was defined as:

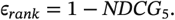

Lower ranking error indicated that higher observed-utility actions were placed near the top of the ranked list. Top-1 regret measured the utility loss from the first-ranked action:

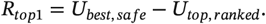

Here, *U*_*best, safe*_ is the observed utility of the best feasible safe action in the portfolio, and *U*_*top, ranked*_ is the observed utility of the model’s first-ranked action. The sweet spot in Figure 3F therefore represents systems that placed high-utility actions near the top and selected a strong first action.

#### EIG calibration evaluation

Figure 3G evaluated whether predicted EIG matched observed information gain after action execution. For each model, candidate actions were assigned a pre-execution predicted EIG. The action was then evaluated through the same execution-and-scoring pipeline to obtain observed information gain. Calibration was assessed by plotting predicted EIG against observed information gain. The identity line represented perfect calibration.

Calibration error was quantified as the absolute difference between predicted and observed EIG:

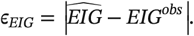

Lower calibration error indicated better prospective estimation of action value. In Figure 3G, the fitted trend for MedGenesis was closest to the identity line, indicating that its predicted action priority better matched realized information gain after execution.

### Real-world clinical investigations

#### Meta-analysis of ICI therapy in cancer patients with CKD

For the L1 evidence-synthesis task, MedGenesis was asked whether CKD modifies the survival benefit of ICI therapy in cancer patients. It executed a PRISMA-compliant systematic review and meta-analysis. PubMed, Embase, and the Cochrane Library were searched through 2024 using Boolean queries covering ICI agents, CKD terms, renal-function strata, and survival outcomes. Eligibility required a cancer cohort with ICI exposure or comparison and an extractable HR for overall survival. Title and abstract screening, full-text review, and structured data extraction were performed sequentially, with uncertain records flagged for human review.

From 2,420 screened records, 15 studies were retained for qualitative synthesis, and 3 studies contributed quantitatively comparable overall-survival HRs for pooling. Log HRs and standard errors were extracted or reconstructed from reported CIs. Pooled estimates were calculated with a random-effects model using REML. Heterogeneity was quantified with I^2^. CKD severity was analyzed in prespecified strata when reported. Subgroups supported by a single study or by wide CIs were treated as exploratory and were not used for causal or practice-changing inference.

#### CARES-009 PTSI pretreatment prognostic stratifier

For the L2 trial-enrichment task, MedGenesis analyzed the single-center Zhongshan Hospital subset of the multicenter CARES-009 perioperative HCC trial (NCT04521153). The analytic dataset included 106 patients in the perioperative treatment arm and 124 patients in the surgery-alone arm. Pretreatment variables included tumor markers, hematologic indices, liver-function tests, and baseline clinical characteristics. EFS was defined from treatment or surgery initiation to recurrence, progression, death, or last follow-up according to the trial endpoint definition.

Within the perioperative arm, MedGenesis first called ViCTOR to contrast patient-state embeddings between recurrent and non-recurrent patients. Feature-attribution analysis identified PIVKA-II, TBIL, SII (platelet count × neutrophil count / lymphocyte count), and AFP as high-contribution variables, with PIVKA-II, TBIL, and SII forming the most compact and clinically interpretable composite. MedGenesis then constructed the PTSI:

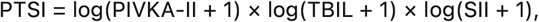

where SII = platelet count × neutrophil count / lymphocyte count. Candidate biomarkers and composite indices were compared by receiver operating-characteristic analysis for recurrence. The PTSI cutoff was selected by the Youden index. Patients were then stratified into low- and high-PTSI groups, and EFS was compared with Kaplan-Meier curves and log-rank tests.

To test whether the recurrence-associated phenotype could support virtual enrollment, low-PTSI perioperative patients were matched 1:1 to patients in the surgery-alone arm by propensity-score matching. Matching variables included log(SII + 1), log(PIVKA-II + 1), and log(AFP + 1), with balance assessed using standardized mean differences. A standardized mean difference below 0.1 was considered acceptable balance. This analysis was treated as hypothesis-generating because the stratifier was derived and tested in a single-center subset; external validation and formal treatment-interaction testing are required before clinical use.

#### GLP-1RA real-world disease-trajectory analysis

For the GLP-1RA application, MedGenesis called ViCTOR to convert longitudinal EHR snapshots from GLP-1RA-exposed patients into patient-state embeddings. The GLP-1RA index date was defined as the first recorded GLP-1RA exposure. Index embeddings from 1,014 exposed patients were projected into principal-component space and grouped into three ViCTOR-derived patient-state phenotypes. Phenotype names were assigned after reviewing state-level enrichment of EHR-derived clinical descriptors.

For short-term trajectory analysis, patients with at least 90 days of post-index follow-up were re-embedded at day 90 after GLP-1RA initiation. Embedding drift was calculated as the Euclidean distance between the index and day-90 ViCTOR embeddings. Paired drift was summarized within each phenotype.

To identify clinical features associated with early embedding drift, laboratory measurements within 90 days before and 90 days after GLP-1RA initiation were paired at the patient level. For each laboratory feature, pre-index and post-index median values were robustly scaled, and absolute paired change was calculated. A feature-driver score was defined as the median absolute standardized paired change multiplied by the absolute Spearman correlation between feature-change magnitude and ViCTOR embedding drift. Features with at least eight paired measurements were ranked and visualized.

To explain the clinical meaning of each ViCTOR phenotype, MedGenesis extracted EHR-derived descriptors including demographics, diagnosis flags, laboratory values, medication orders, procedures, imaging/exam events, and care-intensity measures. Continuous variables were robustly standardized, binary variables were centered by cohort prevalence, and state-level enrichment was calculated for each descriptor. The resulting enrichment matrix was visualized as a phenotype-explanation heatmap and used to annotate the three ViCTOR-derived patient-state phenotypes.

#### Severe immune-related adverse event feature discovery

For the L4 safety-risk task, MedGenesis analyzed pretreatment features associated with severe irAE in ICI-treated HCC patients. The source population contained 6,994 ICI-treated HCC patients. After requiring at least 12 months of outcome follow-up and complete baseline features, 1,120 patients were retained for analysis. Severe irAE was defined as grade 3 or higher toxicity. The final case-control analysis included 101 severe-irAE cases and 404 matched controls.

Rather than using a black-box irAE model score, MedGenesis called ViCTOR for feature attribution on the pretreatment patient-state embeddings. Thirty-one candidate baseline features were screened, including tumor burden, macrovascular invasion, PVTT, liver-function indices, inflammatory markers, autoimmune history, prior treatment, and baseline blood counts. Benjamini-Hochberg FDR control was applied to the screening step. ViCTOR-assisted attribution and regression screening prioritized PVTT, hypoalbuminemia, autoimmune history, and elevated NLR, with lymphocyte count treated as a collinear protective feature rather than an additional score component.

Cases were matched 1:4 to controls using propensity-score matching on age, sex, and baseline clinical covariates. A multivariable conditional-logistic model was then fitted in the matched cohort. Discrimination was assessed by AUC and optimism-corrected C-index. Calibration was assessed by bootstrap resampling and calibration slope. A simplified four-variable screening score was generated by rounding model coefficients: PVTT +1.5, hypoalbuminemia +1.5, autoimmune history +1.0, and elevated NLR +1.0. Patients were stratified into low-, intermediate-, and high-risk groups, and grade 3 or higher irAE incidence was compared across strata using Wilson CIs.

#### Wilson-disease HCC rare-case retrieval and case construction

For the L5 rare-case task, MedGenesis was asked to identify and summarize a rare HCC phenotype arising in Wilson-disease-associated cirrhosis without viral hepatitis. ViCTOR embeddings were generated for 43,812 institutional HCC patients. The query phenotype combined Wilson disease, longterm penicillamine treatment, portal hypertension, absence of viral hepatitis, and subsequent HCC. Candidate patients were ranked by embedding similarity and evidence completeness. The target Wilson-disease HCC case ranked first in the retrieval index.

MedGenesis then assembled a structured case report from the institutional EHR, pathology archive, procedure records, treatment history, and follow-up registry. The output included an 18-year clinical timeline and a 12-marker immunohistochemical panel from the resected tumor, including GPC3, AFP, glutamine synthetase, arginase-1, ARID1α, CK19, CK7, Ki-67, p53, CD34, CD56, and HepPar-1. The final case summary was reviewed by the treating clinical team for factual accuracy and completeness.

##### UK Biobank metabolite prioritization and 3-HB hypothesis generation

For the wet-lab-coupled discovery branch, MedGenesis interrogated the UK Biobank metabolite panel and the published literature in parallel. Each of 249 circulating metabolites was scored for epidemiologic association with incident cancer and for mechanistic coverage in prior literature. The system then used the closed-loop action-priority function to identify candidates with high expected information gain rather than simply ranking by association strength. The two evidence streams converged on ketone-related candidates, and 3-HB was prioritized because the population association was weak and non-linear while the mechanistic literature contained contradictory tumor-suppression and tumor-promotion reports.

##### Public tumor-metabolism and multi-omics immune-context analysis

To place the 3-HB hypothesis in tumor and immune context, MedGenesis queried public tumor-metabolism and multiomics resources, including TCGA, GEO, GTEx, CPTAC, and the Cancer Atlas of Metabolic Profiles^48^. The Cancer Atlas analysis comprised 988 specimens across 11 cancer types and used a weighted consensus across seven immune-deconvolution methods. Associations between 3-HB exposure and inferred immune-cell abundance were compared across B cells, basophils, dendritic cells, eosinophils, macrophages, mast cells, monocytes, natural killer cells, neutrophils, and T cells. Neutrophils showed the most consistent 3-HB-associated increase and were therefore prioritized for tissue and functional validation.

##### Wet-lab-coupled experimental design

MedGenesis first nominated 3-HB as a context-dependent metabolite candidate and then selected the next validation actions in sequence: tumor-cell dose-response testing, syngeneic mouse tumor testing, LY6G immunohistochemistry, human peripheral-blood neutrophil RNA sequencing, and neutrophil-tumor co-culture sufficiency assays. Domain investigators performed all animal, primary-cell, sequencing, staining, and flow-cytometry experiments. Experimental read-outs were returned to MedGenesis as aggregate observations for hypothesis refinement; patient-level and donor-identifying information was not passed to the agent layer.

### Cell culture

Mouse Hepa1-6 hepatoma cells, mouse MC38 colorectal carcinoma cells, human Huh7 hepatocellular carcinoma cells, and human HCCLM3 (LM3) hepatocellular carcinoma cells were cultured under standard mammalian cell-culture conditions. Unless otherwise stated, cells were maintained in high-glucose Dulbecco’s modified Eagle medium (DMEM; Gibco, 11965092) supplemented with 10% fetal bovine serum (FBS; Gibco, 10099141) and 1% penicillin-streptomycin (Gibco, 15140122) at 37°C in a humidified incubator containing 5% CO2. Adherent cells were passaged with 0.25% trypsin-EDTA (Gibco, 25200072) before reaching overconfluence. Cells used for dose-response, implantation, or co-culture assays were in logarithmic growth phase and were confirmed to be mycoplasma-free before use.

### Tumor-cell dose-response assays

To determine whether 3-HB directly affects tumor-cell viability, Hepa1-6, MC38, Huh7, and LM3 cells were seeded in 96-well plates at 2 × 10^3^ cells per well in complete medium. After overnight attachment, cells were treated with sodium 3-hydroxybutyrate (Sigma-Aldrich, 54965) freshly dissolved in phosphate-buffered saline (PBS; Gibco, 10010023), pH-adjusted to 7.4, and sterile-filtered. Serial concentrations of 3-HB were tested at 0, 0.01, 0.1, 1, 10, 100, and 1,000 mmol/L. At 24 h and 48 h after treatment, 10 µL of Cell Counting Kit-8 reagent (CCK-8; Beyotime, C0038) was added to each well and incubated for 2 h at 37°C. Absorbance was measured at 450 nm using a microplate reader. Cell survival was normalized to PBS-treated control wells, and half-maximal inhibitory concentration (IC50) values were estimated by nonlinear regression in GraphPad Prism.

### Mouse tumor models and treatment

Six-week-old male C57BL/6 mice were housed under specific pathogen-free conditions with controlled temperature and humidity and a 12-h light/dark cycle. Hepa1-6 or MC38 cells were harvested during logarithmic growth, washed twice with sterile PBS, and resuspended as single-cell suspensions. For each model, 1 × 10^6^ cells in 100 µL sterile PBS were injected subcutaneously into the right flank. When tumors reached approximately 100 mm^3^, mice were randomized to receive intratumoral PBS vehicle or 3-HB. For each tumor model, 10 mice were assigned to the PBS group and 10 mice were assigned to the 3-HB group. Sodium 3-HB was dissolved in sterile PBS and administered intratumorally at 30 mmol/L in a 100-µL injection volume every 2 days until the prespecified endpoint. Tumor length and width were measured with calipers every 2-3 days, and tumor volume was calculated as length × width^2^ / 2. Mice were euthanized at the endpoint or when humane endpoints were met. Tumors were excised, photographed, and processed for formalin fixation, paraffin embedding, and immunohistochemistry.

### LY6G immunohistochemistry and immune-infiltration quantification

Formalin-fixed, paraffin-embedded Hepa1-6 and MC38 tumor blocks from PBS- and 3-HB-treated mice were sectioned at 4 µm. Sections were deparaffinized in xylene, rehydrated through graded ethanol, and subjected to heat-mediated antigen retrieval in citrate antigen-retrieval buffer, pH 6.0 (Servicebio, G1202), for 15 min. Endogenous peroxidase activity was quenched with 3% hydrogen peroxide for 10 min, followed by blocking with normal goat serum. Sections were incubated overnight at 4°C with anti-LY6G antibody (Abcam, ab210204; clone 1A8; 1:200). Slides were then incubated with horseradish-peroxidase-conjugated goat anti-rat IgG secondary antibody (Abcam, ab97057; 1:500), developed with 3,3’-diaminobenzidine substrate (DAB; ZSGB-BIO, ZLI-9018), counterstained with hematoxylin, dehydrated, and mounted. Digital images were acquired under matched exposure settings. LY6G-positive infiltration was quantified in viable tumor regions by blinded image analysis using ImageJ or QuPath. For each tumor, at least five non-overlapping high-power fields or the whole viable tumor area were analyzed. LY6G abundance was reported as positive area fraction, positive-cell density, or H-score according to the figure panel.

### Human neutrophil isolation and 3-HB treatment

Fresh anticoagulated peripheral blood was collected from healthy human donors and processed within 2 h of collection. Human neutrophils were isolated using either the EasySep Direct Human Neutrophil Isolation Kit (STEMCELL Technologies, 19666) by immunomagnetic negative selection or the human peripheral-blood neutrophil separation kit (Solarbio, P9040) according to the manufacturer’s instructions, with the same isolation method kept consistent within each experimental batch. Residual red blood cells were removed when necessary by brief red-blood-cell lysis, and cells were washed in PBS before culture. Neutrophil purity and viability were assessed by flow cytometry using CD45 and CD66b staining. For RNA-seq and functional stimulation, isolated neutrophils were resuspended in complete RPMI-1640 or DMEM-based medium supplemented with 10% FBS and treated with PBS vehicle or 10 mmol/L 3-HB for 6 h at 37°C and 5% CO2. Matched PBS-treated neutrophils from the same donor were used as controls.

### Human neutrophil RNA sequencing

After PBS or 3-HB treatment, neutrophils were harvested and washed with cold PBS. Total RNA was extracted using TRIzol reagent (Invitrogen, 15596026) followed by column cleanup with the RNeasy Mini Kit (Qiagen, 74104), or an equivalent low-input RNA workflow. RNA quantity and quality were assessed by Qubit fluorometry and Agilent Bioanalyzer. Libraries were prepared using the NEBNext Ultra II Directional RNA Library Prep Kit for Illumina (New England Biolabs, E7760) after poly(A) selection or rRNA depletion according to sample quality. Libraries were sequenced on an Illumina NovaSeq platform using paired-end reads. The primary comparison was 3-HB-treated neutrophils versus paired PBS-treated donor controls.

### Neutrophil RNA-seq processing and gene-set analysis

RNA-seq reads were first inspected with FastQC. Clean reads were aligned to the human reference genome GRCh38 using STAR, and gene-level counts were generated with featureCounts. Differential expression between 3-HB-treated and PBS-treated neutrophils was performed with DESeq2 using donor identity as a blocking factor when paired samples were available. Genes were ranked by signed test statistic or log2 fold change for enrichment analysis. Gene Ontology Biological Process enrichment and gene-set enrichment analysis were performed using clusterProfiler and the Molecular Signatures Database. Chemotaxis, granulocyte/neutrophil migration, leukocyte-mediated cytotoxicity, cell killing, and related effector-function gene sets were prioritized because they were selected by MedGenesis as the discriminating functional axes after the in vivo neutrophil-infiltration result.

### Neutrophil-tumor co-culture sufficiency assays

To test whether 3-HB-conditioned neutrophils were sufficient to promote tumor-cell killing, Huh7 cells were used as target cells in a four-condition factorial co-culture design: Huh7 alone, Huh7 + 3-HB, Huh7 + neutrophils, and Huh7 + neutrophils + 3-HB. Before co-culture, Huh7 cells were seeded in 24-well or 96-well plates and allowed to attach overnight. Freshly isolated human neutrophils were then added at neutrophil-to-tumor ratios of 1:10, 1:1, or 10:1 in the presence or absence of 10 mmol/L 3-HB. Co-cultures were maintained at 37°C with 5% CO2. Reactive oxygen species (ROS) were measured after short-term co-culture using the DCFH-DA-based Reactive Oxygen Species Assay Kit (Beyotime, S0033S), and tumor-cell death was assessed by propidium iodide (PI; BioLegend, 421301) staining. Huh7 + 3-HB and Huh7 + neutrophil conditions served as metabolite-alone and effector-alone controls, respectively.

### Flow cytometry

Flow cytometry was used to assess neutrophil purity, ROS production, and PI-positive tumor-cell death in co-culture assays. Cells were collected, washed with FACS buffer, and stained with fluorochrome-conjugated antibodies for 15-30 min at 4°C in the dark. Human neutrophils were identified as CD45-positive and CD66b-positive cells using APC anti-human CD45 antibody (BioLegend, 304012; clone HI30) and PE or PerCP/Cyanine5.5 anti-human CD66b antibody (BioLegend, 305105 or 305108; clone G10F5). In co-culture experiments, tumor cells were identified as CD45-negative events, while neutrophils were identified as CD45-positive/CD66b-positive events. ROS intensity was quantified as DCF fluorescence intensity in neutrophil-containing conditions. PI-positive killing was quantified as the percentage of PI-positive tumor-cell events among CellTrace-positive/CD45-negative target cells. Data were acquired on a BD LSRFortessa or equivalent flow cytometer and analyzed using FlowJo. Compensation was performed with single-stained controls, and identical gating thresholds were applied across all treatment groups within each experiment.

## QUANTIFICATION AND STATISTICAL ANALYSIS

### Statistical analysis

We defined statistical significance as P < 0.05 and performed statistical analyses using Python (V3.11.2), VSCode, GraphPad Prism (V9.0.0), and the indicated Python packages. Group comparisons were conducted using Student’s t tests, Wilcoxon rank-sum tests, chi-square tests, Fisher’s exact tests, or ANOVA, while paired t tests or paired Wilcoxon tests were used for paired comparisons. Unless otherwise specified, bar plots are presented as mean ± standard deviation (SD), and the exact n values, numbers of biologically independent replicates, and statistical tests are reported in the corresponding figure legends. Correlation analyses used Pearson r or Spearman rho. Survival analyses were performed using Kaplan-Meier curves, log-rank tests, and Cox proportional hazards models, with cutoffs determined by the Youden index or the survminer package where indicated. Meta-analysis, receiver operating characteristic analysis, propensity-score matching, logistic regression, conditional-logistic regression, and multiple-testing correction were performed as described in the corresponding Method details and figure legends. All P values were two-sided unless otherwise specified.

## Abbreviations

3-HB: 3-hydroxybutyrate
AFP: alpha-fetoprotein
AI: artificial intelligence
AUC: area under the curve
CI: confidence interval
CKD: chronic kidney disease
EFS: event-free survival
EHR: electronic health record
EIG: expected information gain
FDR: false discovery rate
GLP-1RA: glucagon-like peptide-1 receptor agonist
HCC: hepatocellular carcinoma
HR: hazard ratio
ICI: immune checkpoint inhibitor
irAE: immune-related adverse event
LLM: large language model
NLR: neutrophil-to-lymphocyte ratio
PBS: phosphate-buffered saline
PI: propidium iodide
PIVKA-II: protein induced by vitamin K absence or antagonist-II
PTSI: pretreatment severity index
PVTT: portal vein tumor thrombus
RNA-seq: RNA sequencing
ROC: receiver operating characteristic
ROS: reactive oxygen species
SII: systemic immune-inflammation index
TBIL: total bilirubin
TCGA: The Cancer Genome Atlas
UR: uncertainty reduction
ViCTOR: Virtual Clinical Trajectory and Observation Representation.

---

**Figure S1.**
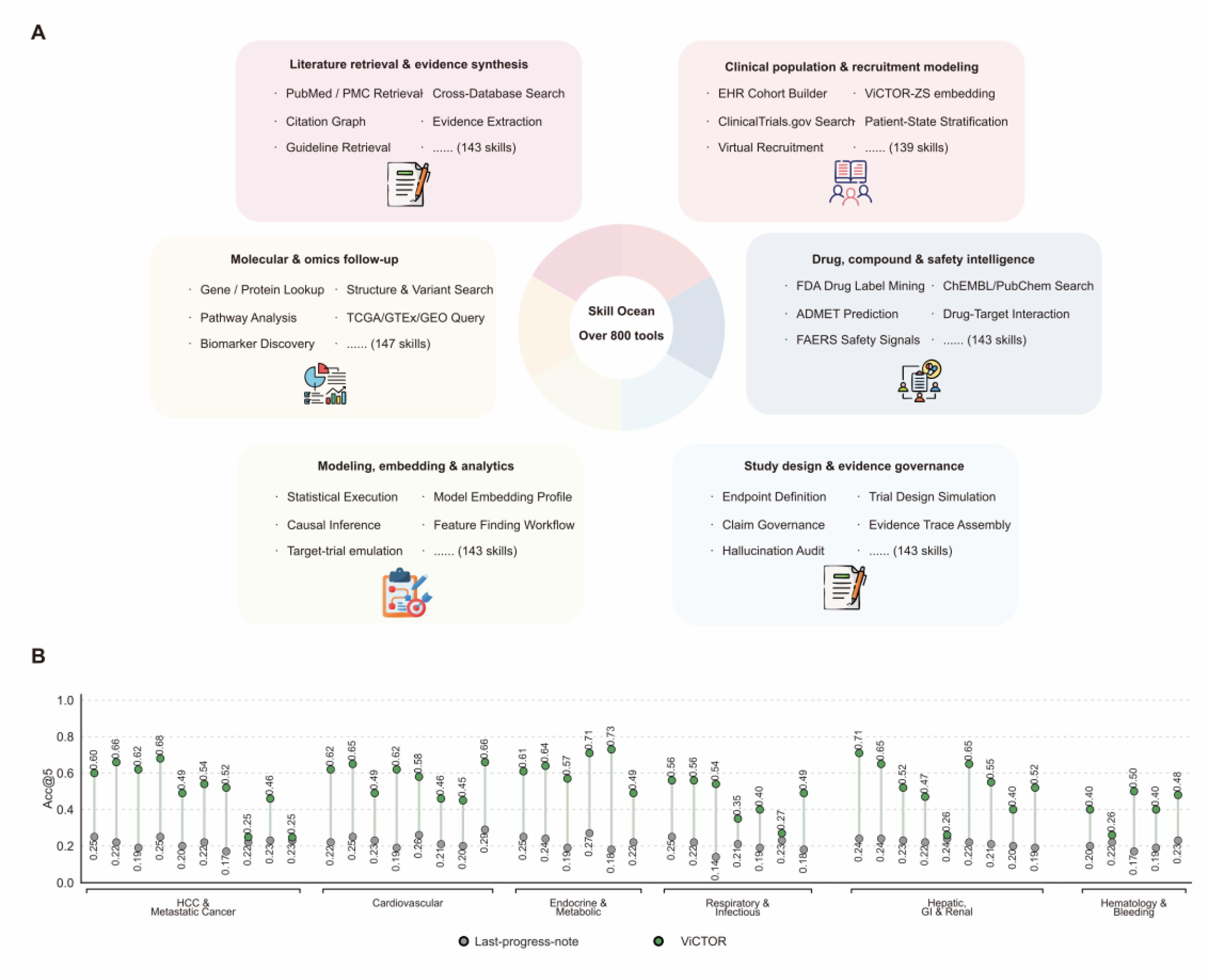
The MedGenesis Skill Ocean and ViCTOR patient-state modeling, related to Figure 1. (A) Skill Ocean overview. MedGenesis uses more than 800 callable tools across six functional families: literature retrieval and evidence synthesis; clinical population and recruitment modeling; molecular and omics followup; drug, compound, and safety intelligence; modeling, embedding, and analytics; and study design and evidence governance. Representative tools and family-level counts are shown. (B) Disease-trajectory retrieval benchmark. Accuracy at 5 (Acc@5) is shown for ViCTOR and the last-progress-note retrieval baseline across HCC and metastatic cancer, cardiovascular disease, endocrine and metabolic disease, respiratory and infectious disease, hepatic/gastrointestinal/renal disease, and hematology or bleeding disorders. Accuracy values are presented as Acc@5.

**Figure S2.**
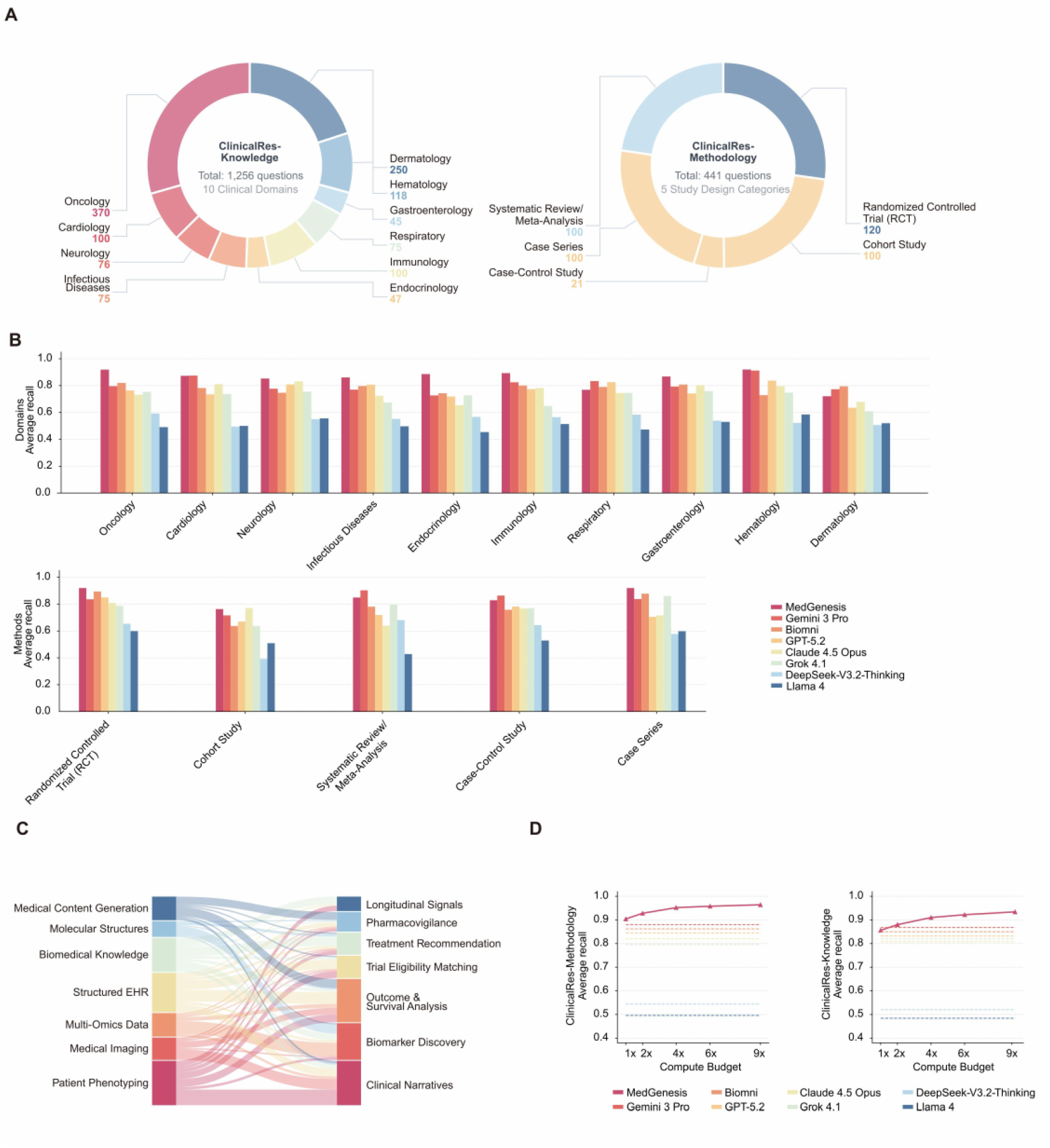
Benchmark composition and scaling behavior, related to Figure 2. (A) Composition of ClinicalResBench. ClinicalRes-Knowledge contains 1,256 questions across 10 clinical domains, and ClinicalRes-Methodology contains 441 questions across 5 study-design categories. (B) Per-domain and per-methodology average recall on ClinicalResBench for MedGenesis and baseline systems. Domains include oncology, cardiology, neurology, infectious diseases, endocrinology, immunology, respiratory medicine, gastroenterology, hematology, and dermatology; methods include randomized controlled trials, cohort studies, systematic review/meta-analysis, case-control studies, and case series. (C) Sankey diagram linking input modalities to downstream clinical-research task types. Input modalities include patient phenotyping, medical imaging, multi-omics data, structured EHR, biomedical knowledge, molecular structures, and medical content generation. Downstream tasks include clinical narratives, biomarker discovery, outcome and survival analysis, trial eligibility matching, treatment recommendation, pharmacovigilance, and longitudinal signal interpretation. (D) Compute-budget scaling curves on ClinicalRes-Methodology and ClinicalRes-Knowledge. Recall is shown from 1x to 9x compute budget for MedGenesis and baseline systems.

**Figure S3.**
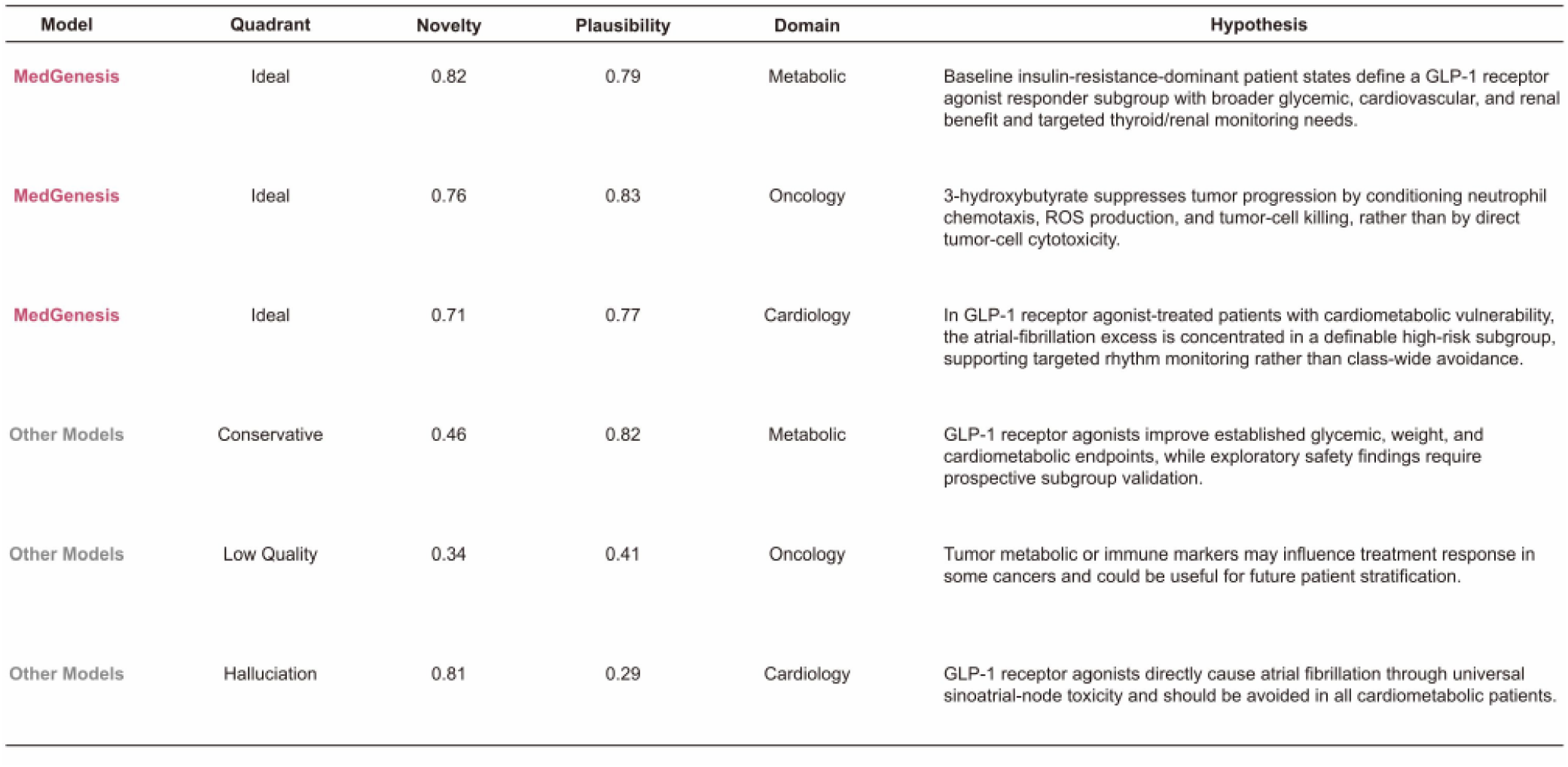
Representative hypothesis quality examples, related to Figure 3. Representative hypotheses generated by MedGenesis and other models under matched metabolic, oncology, and cardiology tasks. Columns show model class, hypothesis-space quadrant, novelty, plausibility, clinical domain, and hypothesis text. MedGenesis examples fall in the Ideal quadrant, whereas baseline examples include conservative, low-quality, and hallucination-prone claims. Novelty and plausibility examples are presented as rubric scores on a 0-1 scale.

**Figure S4.**
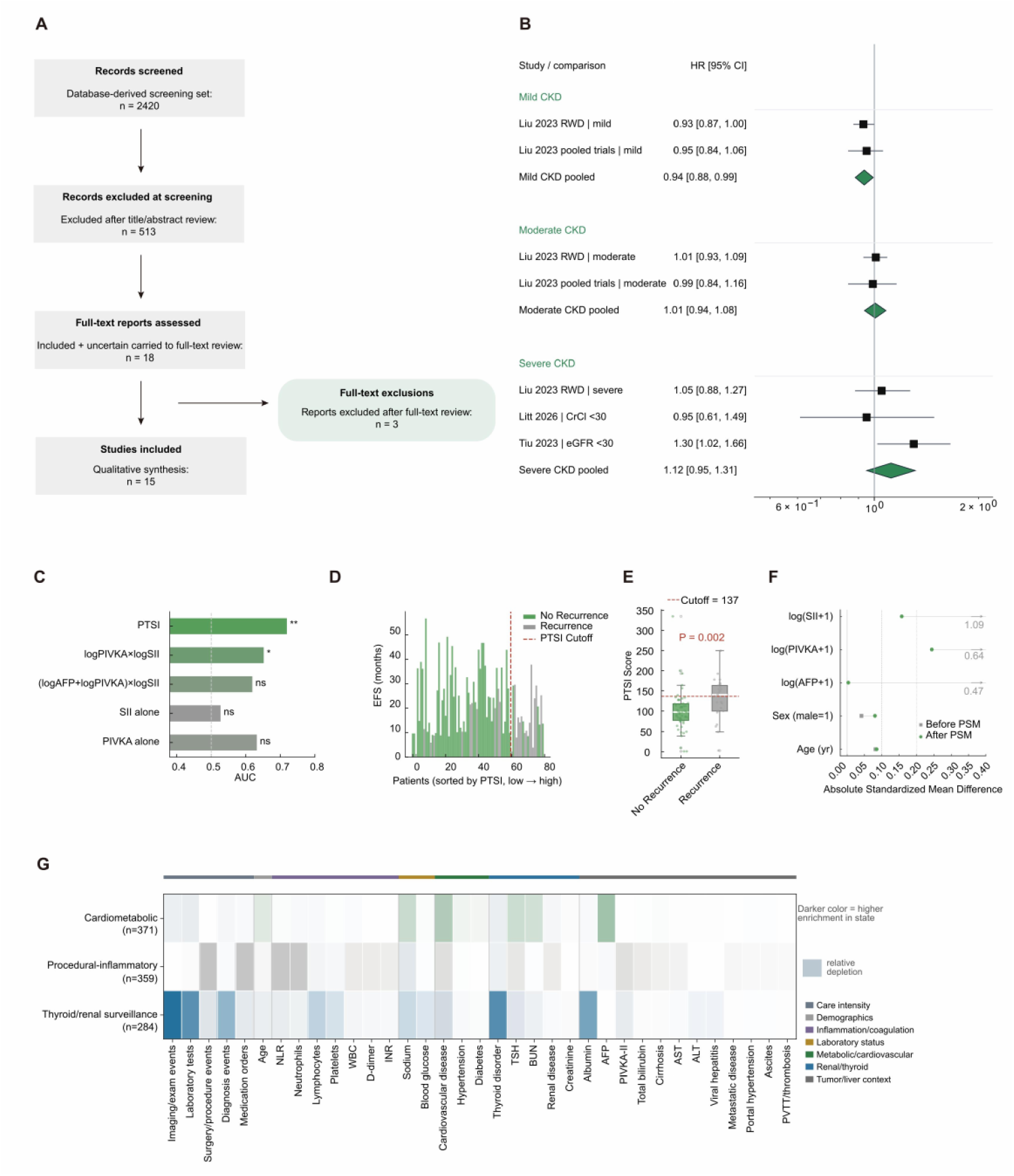
Supplementary outputs for the ICI therapy meta-analysis in CKD and the CARES-009 PTSI stratifier, related to Figure 4. (A) PRISMA-style flow diagram for the ICI therapy in CKD meta-analysis. MedGenesis screened 2,420 database-derived records, excluded 513 after title/abstract review, assessed 18 full-text reports, excluded 3 reports after full-text review, and retained 15 studies for qualitative synthesis. (B) CKD-severity-stratified forest plot. Mild CKD pooled HR = 0.94 (95% CI, 0.88-0.99), moderate CKD pooled HR = 1.01 (95% CI, 0.94-1.08), and severe CKD pooled HR = 1.12 (95% CI, 0.95-1.31). (C) AUC comparison of candidate recurrence-stratification biomarkers and formulas in the CARES-009 Zhongshan Hospital subset. PTSI achieved the highest AUC. (D) PTSI-sorted event-free survival plot in CARES-009 perioperative-arm patients. Bars represent individual patients sorted from low to high PTSI; color denotes recurrence status; the dashed line marks the PTSI cutoff. (E) PTSI score distribution in patients with versus without recurrence. The dashed line marks the cutoff of 137. (F) Covariate balance before and after 1:1 propensity-score matching of low-PTSI perioperative patients against surgery-alone controls. Absolute standardized mean differences are shown for age, sex, log(AFP + 1), log(PIVKA-II + 1), and log(SII + 1). (G) Phenotype-explanation heatmap for the GLP-1RA trajectory analysis. Rows show the three ViCTOR-derived trajectory phenotypes and columns show clinical descriptors grouped by care intensity, demographics, inflammation and coagulation, laboratory status, metabolic/cardiovascular status, renal/thyroid surveillance, and tumor/liver context; darker color indicates higher enrichment in state. Data in the bar plot are presented as AUC values (C), and PTSI distributions are presented as median with interquartile range (E). Effect estimates in forest plots are presented as hazard ratios with 95% confidence intervals (B). P values are displayed directly in the figure where shown, or denoted as ns, not significant; *, P < 0.05; **, P < 0.01. Statistical analyses include random-effects REML meta-analysis (B), DeLong test (C), Mann-Whitney U test (E), and standardized mean-difference assessment after propensity-score matching (F).

**Figure S5.**
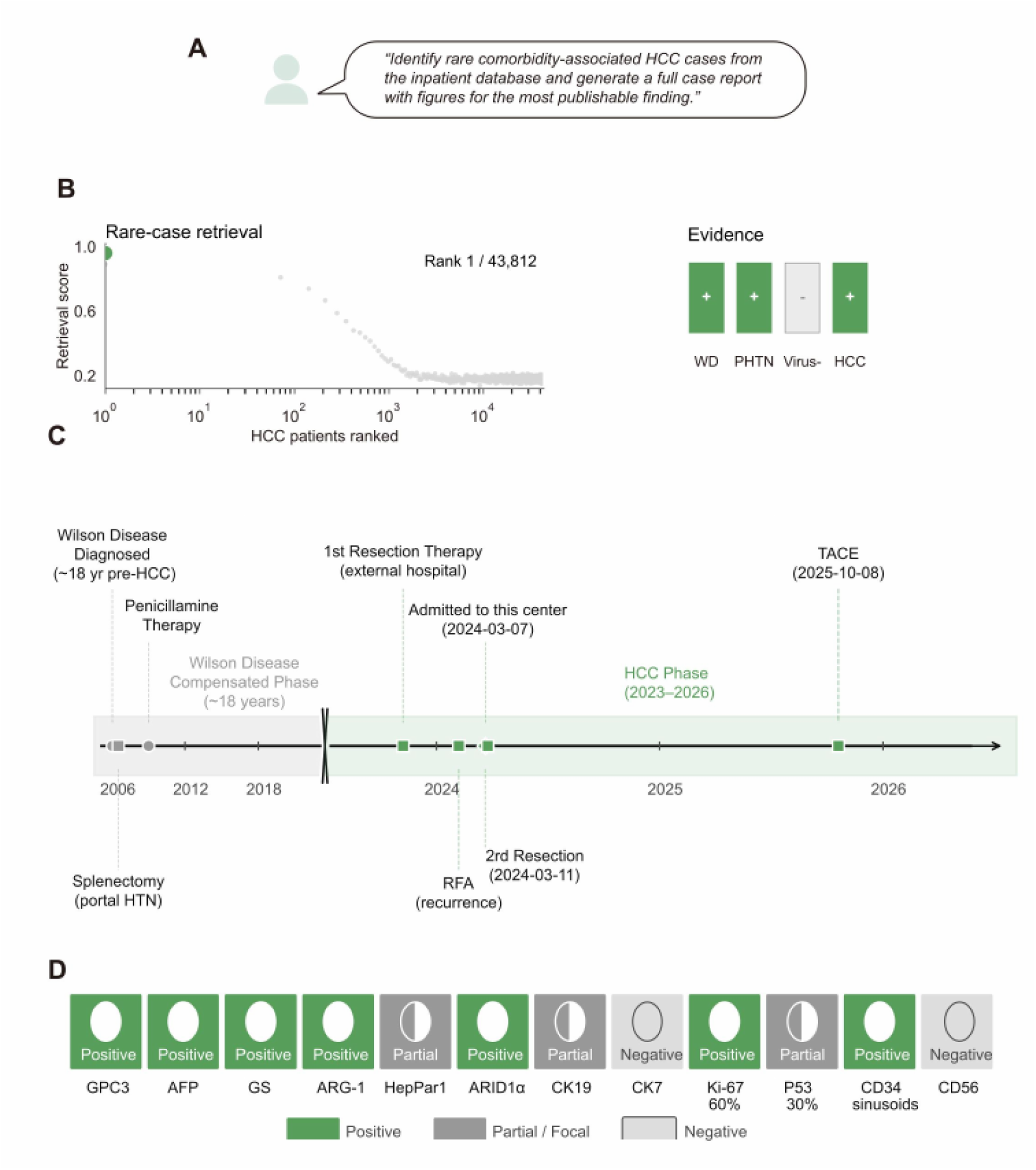
Wilson-disease HCC case retrieval, related to Figure 4. (A) User prompt requesting rare comorbidity-associated HCC case identification and report generation. (B) ViCTOR rare-case retrieval across 43,812 institutional HCC patients. The Wilson-disease-associated, viral-hepatitis-negative HCC case ranked first. The evidence barcode summarizes Wilson disease, portal hypertension, viral-hepatitis-negative status, and HCC evidence. (C) Eighteen-year clinical timeline of the Wilson-disease HCC case, including Wilson disease diagnosis, penicillamine therapy for Wilson disease, portal-hypertension surgery, HCC phase, resections, radiofrequency ablation (RFA), and transarterial chemoembolization (TACE). (D) Immunohistochemical profile of the resected HCC. The 12-marker panel includes GPC3, AFP, glutamine synthetase (GS), arginase-1 (ARG-1), HepPar-1, ARID1alpha, CK19, CK7, Ki-67, p53, CD34, and CD56.

**Figure S6.**
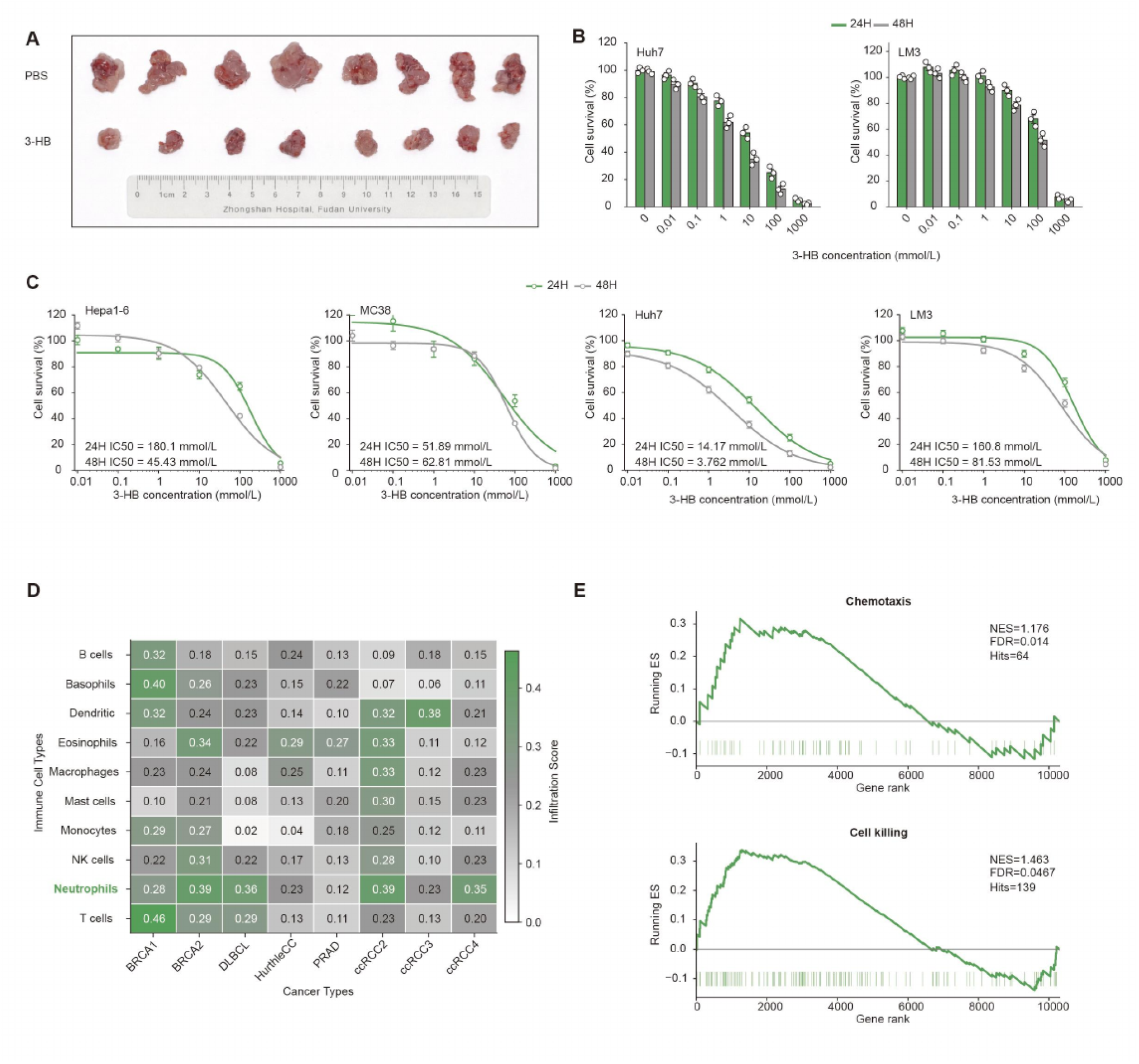
Supplementary mechanistic-validation outputs for the 3-HB-neutrophil axis, related to Figure 6. (A) Excised tumors at endpoint from the in vivo 3-HB experiment, comparing PBS-treated and 3-HB-treated groups. (B) Cell-survival dose-response assays in the human HCC cell lines Huh7 and HCCLM3 (LM3) across serial 3-HB concentrations at 24 h and 48 h. (C) Four-cell-line IC50 curves for 3-HB in Hepa1-6, MC38, Huh7, and HCCLM3 (LM3) cells. 24-h IC50 values were 180.1 mmol/L for Hepa1-6, 51.89 mmol/L for MC38, 14.17 mmol/L for Huh7, and 160.8 mmol/L for HCCLM3. 48-h IC50 values were 45.43 mmol/L, 62.81 mmol/L, 3.762 mmol/L, and 81.53 mmol/L, respectively. (D) Pan-cancer immune-infiltration heatmap across Cancer Atlas of Metabolic Profiles cancer types. Neutrophils are highlighted and show recurrent high infiltration scores across multiple cancer types. (E) Gene set enrichment analysis (GSEA) plots for chemotaxis and cell-killing programs in independent pan-cancer transcriptomes. Chemotaxis: normalized enrichment score (NES) = 1.176, false discovery rate (FDR) = 0.014, hits = 64. Cell killing: NES = 1.463, FDR = 0.0467, hits = 139. Data in the bar plots and dose-response curves are presented as mean +/-standard deviation (B and C). Immune-infiltration values are displayed as consensus infiltration scores (D). GSEA results are reported as normalized enrichment score and false discovery rate (E); FDR < 0.05 was considered significant.

## REFERENCES

1. Subbiah, V. (2023). The next generation of evidence-based medicine. Nature medicine 29, 49–58. 10.1038/s41591-022-02160-z.

2. Sherman, R.E., Anderson, S.A., Dal Pan, G.J., Gray, G.W., Gross, T., Hunter, N.L., LaVange, L., Marinac-Dabic, D., Marks, P.W., Robb, M.A., et al. (2016). Real-World Evidence - What Is It and What Can It Tell Us? The New England journal of medicine 375, 2293–2297. 10.1056/NEJMsb1609216.

3. Shilo, S., Rossman, H., and Segal, E. (2020). Axes of a revolution: challenges and promises of big data in healthcare. Nature medicine 26, 29–38. 10.1038/s41591-019-0727-5.

4. Zhao, W., Wu, C., Fan, Y., Qiu, P., Zhang, X., Sun, Y., Zhou, X., Zhang, S., Peng, Y., Wang, Y., et al. (2026). An agentic system for rare disease diagnosis with traceable reasoning. Nature 651, 775–784. 10.1038/s41586-025-10097-9.

5. Nori, H., and et al. (2025). Sequential diagnosis with language models. https://arxiv.org/abs/2506.22405.

6. Zhang, Z., and et al. (2025). OriGene: a self-evolving virtual disease biologist automating therapeutic target discovery. 10.1101/2025.06.03.657658.

7. Huang, K., Zhang, S., Wang, H., Qu, Y., Lu, Y., Roohani, Y., Li, R., Qiu, L., Li, G., Zhang, J., et al. (2025). Biomni: A General-Purpose Biomedical AI Agent. bioRxiv : the preprint server for biology. 10.1101/2025.05.30.656746.

8. Gottweis, J., and et al. (2025). Towards an AI co-scientist. https://arxiv.org/abs/2502.18864.

9. Singhal, K., Tu, T., Gottweis, J., Sayres, R., Wulczyn, E., Amin, M., Hou, L., Clark, K., Pfohl, S.R., Cole-Lewis, H., et al. (2025). Toward expert-level medical question answering with large language models. Nature medicine 31, 943–950. 10.1038/s41591-024-03423-7.

10. Tu, T., Schaekermann, M., Palepu, A., Saab, K., Freyberg, J., Tanno, R., Wang, A., Li, B., Amin, M., Cheng, Y., et al. (2025). Towards conversational diagnostic artificial intelligence. Nature 642, 442–450. 10.1038/s41586-025-08866-7.

11. McDuff, D., Schaekermann, M., Tu, T., Palepu, A., Wang, A., Garrison, J., Singhal, K., Sharma, Y., Azizi, S., Kulkarni, K., et al. (2025). Towards accurate differential diagnosis with large language models. Nature 642, 451–457. 10.1038/s41586-025-08869-4.

12. Hager, P., Jungmann, F., Holland, R., Bhagat, K., Hubrecht, I., Knauer, M., Vielhauer, J., Makowski, M., Braren, R., Kaissis, G., and Rueckert, D. (2024). Evaluation and mitigation of the limitations of large language models in clinical decision-making. Nature medicine 30, 2613–2622. 10.1038/s41591-024-03097-1.

13. Zhang, K., Zhou, R., Adhikarla, E., Yan, Z., Liu, Y., Yu, J., Liu, Z., Chen, X., Davison, B.D., Ren, H., et al. (2024). A generalist vision-language foundation model for diverse biomedical tasks. Nature medicine 30, 3129–3141. 10.1038/s41591-024-03185-2.

14. Singhal, K., Azizi, S., Tu, T., Mahdavi, S.S., Wei, J., Chung, H.W., Scales, N., Tanwani, A., Cole-Lewis, H., Pfohl, S., et al. (2023). Large language models encode clinical knowledge. Nature 620, 172–180. 10.1038/s41586-023-06291-2.

15. Pal, A., Umapathi, L.K., and Sankarasubbu, M. (2023). Med-HALT: medical domain hallucination test for large language models. 2023. pp. 314–334.

16. Cancer Genome Atlas Research, N., Weinstein, J.N., Collisson, E.A., Mills, G.B., Shaw, K.R.M., Ozenberger, B.A., Ellrott, K., Shmulevich, I., Sander, C., and Stuart, J.M. (2013). The Cancer Genome Atlas Pan-Cancer analysis project. Nature genetics 45, 1113–1120. 10.1038/ng.2764.

17. Edgar, R., Domrachev, M., and Lash, A.E. (2002). Gene Expression Omnibus: NCBI gene expression and hybridization array data repository. Nucleic acids research 30, 207–210. 10.1093/nar/30.1.207.

18. Consortium, G.T. (2020). The GTEx Consortium atlas of genetic regulatory effects across human tissues. Science (New York, N.Y.) 369, 1318–1330. 10.1126/science.aaz1776.

19. Edwards, N.J., Oberti, M., Thangudu, R.R., Cai, S., McGarvey, P.B., Jacob, S., Madhavan, S., and Ketchum, K.A. (2015). The CPTAC Data Portal: A Resource for Cancer Proteomics Research. Journal of proteome research 14, 2707–2713. 10.1021/pr501254j.

20. Sayers, E.W., Bolton, E.E., Brister, J.R., Canese, K., Chan, J., Comeau, D.C., Connor, R., Funk, K., Kelly, C., Kim, S., et al. (2022). Database resources of the national center for biotechnology information. Nucleic acids research 50, D20–D26. 10.1093/nar/gkab1112.

21. National Cancer, I. (2024). Surveillance, Epidemiology, and End Results (SEER) Program.

22. Centers for Disease, C., and Prevention (2024). National Health and Nutrition Examination Survey (NHANES).

23. Sudlow, C., Gallacher, J., Allen, N., Beral, V., Burton, P., Danesh, J., Downey, P., Elliott, P., Green, J., Landray, M., et al. (2015). UK biobank: an open access resource for identifying the causes of a wide range of complex diseases of middle and old age. PLoS medicine 12, e1001779. 10.1371/journal.pmed.1001779.

24. Johnson, A.E.W., Bulgarelli, L., Shen, L., Gayles, A., Shammout, A., Horng, S., Pollard, T.J., Hao, S., Moody, B., Gow, B., et al. (2023). MIMIC-IV, a freely accessible electronic health record dataset. Scientific data 10, 1. 10.1038/s41597-022-01899-x.

25. Bedi, S., Liu, Y., Orr-Ewing, L., Dash, D., Koyejo, S., Callahan, A., Fries, J.A., Wornow, M., Swaminathan, A., Lehmann, L.S., et al. (2025). Testing and Evaluation of Health Care Applications of Large Language Models: A Systematic Review. JAMA 333, 319–328. 10.1001/jama.2024.21700.

26. Google (2025). A new era of intelligence with Gemini 3. Google Blog.

27. OpenAi (2025). Introducing GPT-5.2. OpenAI.

28. Anthropic (2025). Claude Opus 4.5 system card. Anthropic.

29. xAi (2025). Grok 4.1 model card. xAI.

30. DeepSeek, A.I., Aixin, L., Aoxue, M., Bangcai, L., Bing, X., Bingxuan, W., Bingzheng, X., Bochao, W., Bowei, Z., Chaofan, L., et al. (2025). DeepSeek-V3.2: Pushing the Frontier of Open Large Language Models. https://arxiv.org/abs/2512.02556.

31. Meta, A.I. (2025). The Llama 4 herd: the beginning of a new era of natively multimodal AI innovation. Meta AI.

32. OpenAi (2025). Codex CLI. GitHub repository.

33. Anthropic (2025). Claude Code overview. Anthropic.

34. Hartzell, S., Bin, S., Cantarelli, C., Haverly, M., Manrique, J., Angeletti, A., Manna, G., Murphy, B., Zhang, W., Levitsky, J., et al. (2020). Kidney Failure Associates With T Cell Exhaustion and Imbalanced Follicular Helper T Cells. Front Immunol 11, 583702. 10.3389/fimmu.2020.583702.

35. Espi, M., Koppe, L., Fouque, D., and Thaunat, O. (2020). Chronic Kidney Disease-Associated Immune Dysfunctions: Impact of Protein-Bound Uremic Retention Solutes on Immune Cells. Toxins (Basel) 12. 10.3390/toxins12050300.

36. Liu, Q., Mathur, R., Xu, Y., Torres, A.Z., Miksad, R.A., Liu, C., Smithson, H., Wang, Y., Zhu, H., Booth, B., et al. (2023). The Association Between Baseline Hepatic or Renal Function and Clinical Outcomes for Patients With Non-Small Cell Lung Cancer Treated With a PD-1/PD-L1 Blocking Antibody Using Real-World and Trial Data. Clinical pharmacology and therapeutics 113, 1139–1149. 10.1002/cpt.2874.

37. Tiu, B.C., Strohbehn, I.A., Zhao, S., Ouyang, T., Hanna, P., Wang, Q., Gupta, S., Leaf, D.E., Reynolds, K.L., and Sise, M.E. (2023). Safety of Immune Checkpoint Inhibitors in Patients With Advanced Chronic Kidney Disease: A Retrospective Cohort Study. The oncologist 28, e379–e390. 10.1093/oncolo/oyad001.

38. Wang, Z., Fan, J., Zhou, S., Sun, Y., Liang, F., Ji, Y., Gu, F., Li, T., Peng, L., Peng, T., et al. (2025). Perioperative camrelizumab plus rivoceranib versus surgery alone in patients with resectable hepatocellular carcinoma at intermediate or high risk of recurrence (CARES-009): a randomised phase 2/3 trial. Lancet (London, England) 406, 2089–2099. 10.1016/S0140-6736(25)01720-9.

39. Qin, S., Gu, S., Chan, S.L., Bai, Y., Ren, Z., Lin, X., Chen, Z., Jia, W., Jin, Y., Guo, Y., et al. (2025). Camrelizumab plus rivoceranib versus sorafenib as first-line therapy for unresectable hepatocellular carcinoma (CARES-310): final analysis of a randomised, open-label, international, phase 3 study. The Lancet. Oncology 26, 1598–1611. 10.1016/S1470-2045(25)00543-1.

40. Dai, H., Li, Y., Lee, Y.A., Lu, Y., George, T.J., Donahoo, W.T., Lee, K.P., Nakshatri, H., Allen, J., Guo, Y., et al. (2025). GLP-1 Receptor Agonists and Cancer Risk in Adults With Obesity. JAMA oncology 11, 1186–1193. 10.1001/jamaoncol.2025.2681.

41. Baxter, S.M., Lund, L.C., Andersen, J.H., Brix, T.H., Hegedüs, L., Hsieh, M.H.-C., Su, C.T.-T., Cheng, M.C.-Y., Chang, Z.C.-J., Lai, E.C.-C., et al. (2025). Glucagon-Like Peptide 1 Receptor Agonists and Risk of Thyroid Cancer: An International Multisite Cohort Study. Thyroid : official journal of the American Thyroid Association 35, 69–78. 10.1089/thy.2024.0387.

42. Wang, L., Xu, R., Kaelber, D.C., and Berger, N.A. (2024). Glucagon-Like Peptide 1 Receptor Agonists and 13 Obesity-Associated Cancers in Patients With Type 2 Diabetes. JAMA network open 7, e2421305. 10.1001/jamanetworkopen.2024.21305.

43. Pfeiffenberger, J., Mogler, C., Gotthardt, D.N., Schulze-Bergkamen, H., Litwin, T., Reuner, U., Hefter, H., Huster, D., Schemmer, P., Członkowska, A., et al. (2015). Hepatobiliary malignancies in Wilson disease. Liver Int 35, 1615–1622. 10.1111/liv.12727.

44. van Meer, S., de Man, R.A., van den Berg, A.P., Houwen, R.H., Linn, F.H., van Oijen, M.G., Siersema, P.D., and van Erpecum, K.J. (2015). No increased risk of hepatocellular carcinoma in cirrhosis due to Wilson disease during long-term follow-up. J Gastroenterol Hepatol 30, 535–539. 10.1111/jgh.12716.

45. Reddy, K.R., McLerran, D., Marsh, T., Parikh, N., Roberts, L.R., Schwartz, M., Nguyen, M.H., Befeler, A., Page-Lester, S., Tang, R., et al. (2023). Incidence and Risk Factors for Hepatocellular Carcinoma in Cirrhosis: The Multicenter Hepatocellular Carcinoma Early Detection Strategy (HEDS) Study. Gastroenterology 165, 1053–1063.e1056. 10.1053/j.gastro.2023.06.027.

46. Dang, Q., Li, B., Jin, B., Ye, Z., Lou, X., Wang, T., Wang, Y., Pan, X., Hu, Q., Li, Z., et al. (2024). Cancer immunometabolism: advent, challenges, and perspective. Molecular cancer 23, 72. 10.1186/s12943-024-01981-5.

47. Murphy, S., Rahmy, S., Gan, D., Liu, G., Zhu, Y., Manyak, M., Duong, L., He, J., Schofield, J.H., Schafer, Z.T., et al. (2024). Ketogenic Diet Alters the Epigenetic and Immune Landscape of Prostate Cancer to Overcome Resistance to Immune Checkpoint Blockade Therapy. Cancer research 84, 1597–1612. 10.1158/0008-5472.CAN-23-2742.

48. Benedetti, E., Liu, E.M., Tang, C., Kuo, F., Buyukozkan, M., Park, T., Park, J., Correa, F., Hakimi, A.A., Intlekofer, A.M., et al. (2023). A multimodal atlas of tumour metabolism reveals the architecture of gene-metabolite covariation. Nature metabolism 5, 1029–1044. 10.1038/s42255-023-00817-8.

